# Impact of transnational gentrification on the health of adults from the EPIPorto cohort: a quasi-experimental study

**DOI:** 10.64898/2026.11.08.26367550

**Authors:** José António Figueiredo, Henrique Alves, Carla Moreira, Ana Isabel Ribeiro

**Affiliations:** São João Public Health Unit, São João Local Health Unit, Alameda Professor Hernâni Monteiro, 4200-319 Porto, Portugal; EPIUnit ITR, Instituto de Saúde Pública da Universidade do Porto, Universidade do Porto, Rua das Taipas, n° 135, 4050-600 Porto, Portugal; MEDCIDS - Department of Community Medicine, Information and Health Decision Sciences, Faculty of Medicine, University of Porto, Alameda Professor Hernâni Monteiro, 4200-319 Porto, Portugal; RISE-Health, Faculty of Medicine, University of Porto, Alameda Professor Hernâni Monteiro, 4200-319 Porto, Portugal; CEGOT - Centre of Studies in Geography and Spatial Planning, Faculty of Arts and Humanities of Porto, University of Porto, Via Panorâmica s/n, 4150-564 Porto, Portugal

**Keywords:** Gentrification, Public Health, Quasi-experimental study, Social determinants of health, Urban Health

## Abstract

Gentrification is a global, accelerating urban process involving complex socio-spatial transformations. Although increasingly recognised as a determinant of urban health, quantitative evidence on its population health impacts remains scarce and inconsistent, limiting evidence-informed urban policymaking, especially in rapidly gentrifying contexts. Thus, we assessed the health impact of neighbourhood-level gentrification among adults in Porto (Northern Portugal), a hotspot of transnational gentrification. A quasi-experimental study was conducted using data from the EPIPorto population-based cohort (1999–2022), with 2,485 participants at baseline. A validated, context-sensitive 16-variable index of transnational gentrification, derived from 2011 and 2021 census and tourism-accommodation data through Principal Component Analysis, enabled neighbourhood classification as gentrifying or non-gentrifying. Difference-in-differences models estimated the effect of living in gentrifying neighbourhoods on ten cardiometabolic and behavioural outcomes. Inverse probability of censoring weighting was used to account for loss to follow-up and missing data. At baseline, 26.2% of participants resided in gentrifying neighbourhoods. Residence in a gentrifying neighbourhood was associated with a statistically significant reduction in nightly sleep duration (−0.346 h; 95% CI −0.650, −0.042), with statistically significant reductions in the lower-education and ≥65-year subgroups. No significant effects were observed for body mass index, overweight/obesity, total/HDL/LDL cholesterol, blood pressure, and tobacco or alcohol use. Transnational gentrification in Porto showed limited effects on most cardiometabolic outcomes but was associated with reduced sleep duration, suggesting that the health impacts of gentrification may initially emerge through psychosocial pathways before translating into measurable cardiometabolic changes.

## 1. Introduction

With nearly two-thirds of the world’s population projected to live in urban areas by 2050 (United Nations Department of Economic and Social Affairs, 2018) and up to 80% by 2100 (Jones et al., 2020), cities have become central to contemporary public health (Kadakia C Galea, 2023). Growing attention has therefore been directed towards the urban environment as a determinant of population health and to the neighbourhood-level processes that reshape it (Kadakia C Galea, 2023; Ribeiro, 2018), among which gentrification has become prominent in urban health research (Cole, 2020).

Gentrification is a socio-spatial process in which reinvestment in the built environment to suit the preferences of higher-socioeconomic-status residents and users (Cole, 2020) is typically accompanied by the in-migration of more affluent newcomers, rising housing costs and, frequently, the displacement of long-standing residents (Sigler C Wachsmuth, 2020). By reshaping both the physical and social dimensions of the urban environment (Bryson, 2013) – including the food environment (Breyer C Voss-Andreae, 2013), green spaces (Jelks et al., 2021), social cohesion and safety (Schnake-Mahl, Jahn, et al., 2020) – gentrification can plausibly affect a broad range of health outcomes, acting as a double-edged process that may simultaneously create health-promoting opportunities and undermine health (Schnake-Mahl, Jahn, et al., 2020).

The local food environment and access to health-promoting resources illustrate these opposing influences. Gentrification may improve access to healthier food options and to renovated spaces for physical activity, such as public parks. However, as prices rise and the commercial offer changes, such amenities may become unaffordable to, or poorly aligned with the needs of, long-standing residents, who may also lose established local resources and services that supported their daily routines and health-related behaviours (Breyer C Voss-Andreae, 2013; Schroeder et al., 2019). Displaced residents may not benefit from these improvements at all.

Conceptual frameworks on gentrification and urban health describe several pathways through which neighbourhood change may influence physical and mental health, and emphasise that these effects may be unevenly distributed across residents (Cole et al., 2021; Cole et al., 2024). Through psychosocial and neighbourhood-level mechanisms, gentrification may shape health-related behaviours such as alcohol and tobacco use (Pennay et al., 2014), impair sleep (Bozick et al., 2021) and affect downstream cardiometabolic outcomes such as body weight and blood pressure (Chen et al., 2023; Cole et al., 2021).

Reflecting this complexity, previous studies have reported beneficial, null and harmful health effects of gentrification, and reviews have found no consensus as to whether its net influence on population health is positive or negative (Bhavsar et al., 2020; Delong, 2023; Schnake-Mahl, Jahn, et al., 2020; Tulier et al., 2019). In Los Angeles County, for example, gentrification was associated with modestly better hypertension control, more markedly among non-Hispanic white and older residents, but not with diabetes control (Chen et al., 2023).

In a large prospective cohort of Hispanic/Latino adults, neighbourhood socioeconomic change was not associated with the six-year incidence of metabolic syndrome (Pichardo et al., 2023), whereas adverse associations, including poorer self-rated health and greater psychological distress, have been consistently documented among historically marginalised residents (Anguelovski et al., 2021).

This heterogeneity suggests that the health effects of gentrification depend on individual vulnerability, local context and the specific pathways involved. Reviews and conceptual work indicate that the benefits of neighbourhood upgrading accrue mainly to more privileged groups, while socioeconomically disadvantaged residents are harmed or fail to benefit (Cole et al., 2021; Cole et al., 2024). Age may further shape this vulnerability, as older adults tend to be more exposed to neighbourhood change owing to longer residential tenure, reduced mobility and more constrained financial resources (Torres, 2020). Because beneficial and adverse influences may operate simultaneously within the same population, and even on the same outcome (Delong, 2023), they may offset one another when effects are estimated for residents as a whole, masking both the concentration of harm among the most disadvantaged and the resulting widening of health inequities (Cole et al., 2024).

Part of this inconsistency may also be methodological. Most studies have relied on cross-sectional designs that cannot distinguish the effects of gentrification from pre-existing neighbourhood differences (Delong, 2023), and the operationalisation of gentrification remains heterogeneous (Bhavsar et al., 2020). Because gentrification unfolds gradually and its health effects accrue over time, longitudinal and quasi-experimental designs are better suited to disentangling its impact from underlying temporal trends and confounding (Delong, 2023; Schnake-Mahl, Sommers, et al., 2020; Tulier et al., 2019).

Although gentrification is a global phenomenon, its health impacts remain comparatively understudied in European settings (Delong, 2023; Schnake-Mahl, Sommers, et al., 2020). In the city of Porto, northern Portugal, gentrification has intensified rapidly over the past decade and a half, driven by three interlinked factors: the expansion of Francisco Sá Carneiro Airport from 2007 onwards, increased real-estate investment following the 2008 financial crisis, and growing national and international student inflows (Carvalho et al., 2019). Its transnational character, reflected both in the origin of capital and in the international composition of newcomers (Silva et al., 2023; Sousa C Barcón, 2021), makes Porto a particularly relevant European setting in which to study the health impacts of gentrification (Ribeiro et al., 2025).

In Porto, two recent qualitative studies have documented both perceived benefits (e.g., urban renewal, improved transport) and harms (e.g., insecurity, mental health concerns, reduced social ties) attributed to gentrification (Santos, Silva, et al., 2024; Silva et al., 2023), and a cross-sectional analysis reported an association between housing insecurity and several health and well-being outcomes in older adults (Santos, Henriques, et al., 2024). However, no quantitative study has yet assessed the health impacts of gentrification in Portugal. Against this background, we aimed to estimate the health effects of transnational gentrification in Porto, measured between 2011 and 2021, among adults in the EPIPorto cohort. Secondarily, we assessed whether these effects were modified by educational attainment and age.

## 2. Methods

### 2.1 Study design and participants

A quasi-experimental longitudinal study was conducted within the EPIPorto population-based cohort, which enrolled 2,485 community-dwelling Portuguese adults aged 18 years or older living in Porto between 1999 and 2003 (baseline). Participants were identified through random-digit dialling of household landlines, which covered approximately 97% of households at the time, and one eligible non-institutionalised adult was randomly selected from each household (Ramos et al., 2004). Residences were georeferenced using ArcGIS Online World Geocoding Service and Google Maps, following rigorous geoprivacy-preserving guidelines (Ribeiro, Dias, et al., 2022), enabling spatial linkage with census block group characteristics.

Follow-up assessments were conducted in five waves (Figure 1). As the current gentrification process in Porto was considered to have started around 2013, after the economic crisis (Carvalho et al., 2019), baseline and wave 1 formed the pre-exposure period and subsequent waves the post-exposure period. Wave 2 was excluded from the primary analysis because it occurred early in the gentrification process, when cumulative exposure may not yet have been sufficiently established; its re-inclusion was examined in a sensitivity analysis (Supplementary Material Section E). Wave 4 was not included because none of the outcomes was assessed in that wave.

**Figure 1.**
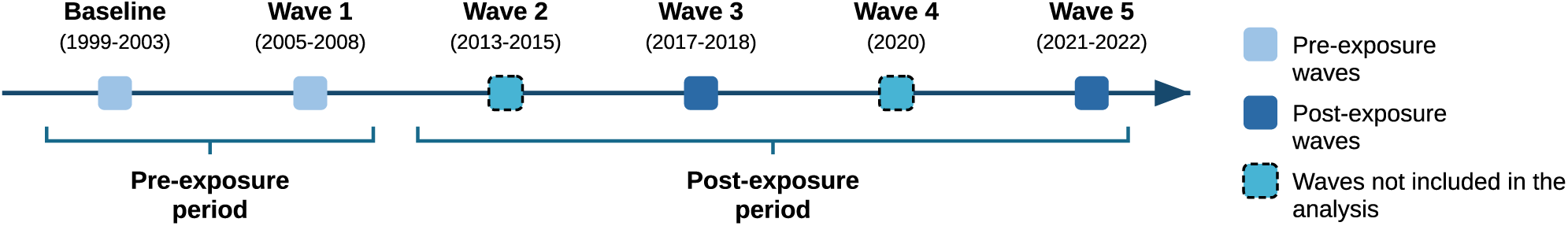
Flow diagram of the EPIPorto cohort showing baseline and the follow-up assessment waves.

All EPIPorto participants were eligible for inclusion. Participants who moved to a census block group with a different gentrification classification during the post-exposure period were excluded from the primary analysis; in a sensitivity analysis, they were retained and classified according to their census block group of residence at wave 3, the first post-exposure wave analysed (Supplementary Material Section G). Participants were allocated to study groups (exposed *vs* unexposed) according to the gentrification status of their census block group of residence (gentrifying *vs* non-gentrifying), and repeated measurements of ten health outcomes obtained between 1999 and 2022 were used to compare outcome trajectories between groups. This study is reported in accordance with the Strengthening the Reporting of Observational Studies in Epidemiology (STROBE) guideline (von Elm et al., 2007).

### 2.2 Exposure assessment

Neighbourhood-level gentrification was operationalised through a multidimensional index derived by Principal Component Analysis (PCA), previously developed and validated for the Porto Metropolitan Area (Alves et al., 2026). The unit of analysis was the census block group, the second smallest geographical unit of the Portuguese census, which in 2021 had, on average, 1,203 residents and an area of 1.41 km² (range 0.03–44.38 km²) in the Porto Metropolitan Area. Indicators were obtained from the 2011 and 2021 national censuses (Instituto Nacional de Estatística [Statistics Portugal, INE]) and the national tourism accommodation register (Turismo de Portugal, I.P. [Tourism of Portugal]).

Fifteen candidate indicators, each expressed as its 2011 value and its absolute change between 2011 and 2021 (30 variables), were considered. Following exploratory analysis of correlations and communalities, 16 standardised (z-score) variables were retained, reflecting both a census block group’s baseline status and its temporal trajectory and covering conceptual domains commonly identified in the literature (Bilal et al., 2020; Cocola-Gant et al., 2020): educational attainment, occupational class, employment status, housing (high rents), demographic composition (older adults, young adults and foreign residents from high-income countries) and tourism (short-term accommodation). The variables included in the index and the spatial distribution of gentrifying census block groups are shown in Supplementary Table S1 and Supplementary Figure S1, respectively.

The principal component selected as the basis of the gentrification index showed high positive loadings for increases in higher-education attainment, foreign residents from high-income countries, young adult population, housing units with high rents, and short-term rentals, alongside a negative loading for change in the proportion of residents aged ≥65 years. It was also effective in distinguishing census block groups with lower socioeconomic status at baseline (Alves et al., 2026), known to be more susceptible to gentrification (Freeman, 2005).

The transnational gentrification index was then standardised (mean=0, SD=1) and dichotomised using a threshold of >1 SD, approximately corresponding to the 90th percentile. Compared with alternative thresholds (+0.5 SD, +2 SD and the 75th, 90th and 95th percentiles), this cut-off showed a strong and consistent association with Property Tax Value, one of the external indicators used to validate the index, while classifying approximately 10% of census block groups as gentrifying (Alves et al., 2026), and most closely matched the spatial distribution of areas previously described as undergoing intense gentrification in Porto (Carvalho et al., 2019; Sousa C Barcón, 2021). Participants residing in census block groups above this threshold (gentrifying) were considered exposed, and those residing in the remaining census block groups (non-gentrifying) unexposed. In sensitivity analyses, gentrifying census block groups were alternatively defined as those scoring ≥1.5 SD above the mean or belonging to the upper quartile of the index (Supplementary Material Section F).

### 2.3 Outcomes

Guided by conceptual frameworks on gentrification and population health (Cole et al., 2021; Cole et al., 2024), ten cardiometabolic and behavioural outcomes with repeated, comparable measurements across follow-up waves were selected. Continuous outcomes were body mass index (BMI, kg/^m²^), total/HDL/LDL cholesterol (mg/dL), systolic and diastolic blood pressure (mmHg), and average nightly sleep duration (hours). Binary outcomes were overweight/obesity, defined as BMI ≥ 25 kg/^m²^ (World Health Organization, 2019), current tobacco use and current alcohol use (Supplementary Material, Section B). BMI, overweight/obesity, total, HDL and LDL cholesterol, and tobacco use were available at baseline and waves 1, 2, 3 and 5; systolic and diastolic blood pressure and alcohol use at baseline and waves 1, 2 and 3; and sleep duration at baseline and waves 1, 3 and 5.

### 2.4 Covariates

Based on previous conceptual frameworks linking gentrification and neighbourhood change to health (Bhavsar et al., 2020; Cole et al., 2021; Cole et al., 2024; Ribeiro, 2022), age, sex, educational attainment, employment status and marital status were treated as potential confounders and included in the adjusted models. All covariates entered the models with their pre-exposure values, so that adjustment was restricted to characteristics that could not have been affected by the exposure. Educational attainment was classified according to the International Standard Classification of Education (ISCED 2011) as low (ISCED 0–2), medium (ISCED 3–4) or high (ISCED 5–8); employment status as employed, unemployed or other (e.g., retired, homemaker or student); and marital status as with or without partner. Covariate categories were harmonised across waves.

Age and educational attainment were additionally examined as potential effect modifiers in stratified analyses. These two roles are not mutually exclusive: adjusting for a variable removes the bias it would otherwise introduce into the overall estimate, whereas examining it as an effect modifier assesses whether the magnitude of that effect differs across its levels (Hernán C Robins, 2020).

Both effect modifiers were selected a priori, given previous evidence that the health impacts of gentrification are unevenly distributed, with benefits accruing mainly to more socioeconomically privileged residents and harms concentrating among socioeconomically disadvantaged and older ones (Cole et al., 2021; Cole et al., 2024). Educational attainment was used as an individual-level marker of socioeconomic position, since lower educational attainment may reflect fewer material and social resources with which to absorb negative consequences of gentrification. Older residents may be particularly susceptible to neighbourhood disruption, owing to longer residential attachment, greater reliance on local social networks and greater baseline health vulnerability (Smith et al., 2018), a pattern also described among older adults in Porto (Santos, Henriques, et al., 2024). Sex was retained as a covariate but was not examined as an effect modifier, as we identified no specific theoretical or empirical basis for expecting differential susceptibility to gentrification-related neighbourhood change by sex, and further stratification would have reduced precision.

### 2.5 Statistical analysis

Each outcome-specific analytical sample was described using means and standard deviations for continuous variables and counts and percentages for categorical variables. Participants living in gentrifying and non-gentrifying census block groups were compared at each wave using Welch’s t-tests and Pearson’s chi-squared tests, as were participants included in and excluded from the complete-case analyses, to assess potential selective loss to follow-up and missingness.

Effects were estimated using the Callaway and Sant’Anna difference-in-differences (DiD) estimator for multiple time periods, which provides the average treatment effect on the treated (ATT) at each post-exposure wave and an aggregated ATT across post-exposure waves, used as the main effect estimate. Compared with traditional two-way fixed-effects models, this estimator accommodates heterogeneous treatment effects, multiple pre- and post-treatment periods, and covariate-conditional assessment of the parallel-trends assumption (Callaway C Sant’Anna, 2021).

Inverse probability of censoring weighting (IPCW) was defined as the primary analytical approach, to reduce potential selection bias arising from loss to follow-up and missing outcome or covariate data, under the assumption that censoring was independent of the potential outcomes conditional on measured covariates. Pooled logistic regression was used to estimate the wave-specific probability of remaining under observation conditional on baseline covariates, and stabilised cumulative weights, truncated at the 1st and 99th percentiles, were incorporated into the DiD models (Cole C Hernán, 2008; Hernán C Robins, 2020; Robins C Finkelstein, 2000).

The parallel-trends assumption, which requires that, in the absence of exposure, outcome trajectories would have evolved similarly in the exposed and unexposed groups, was formally assessed over the pre-exposure period using Wald tests. A non-significant test (p>0.05) was interpreted as consistent with the assumption; otherwise, the corresponding ATT was not interpretable as a causal effect. Complete-case analyses were conducted as a secondary analysis to assess the sensitivity of the findings to the exclusion of participants with incomplete data.

The stability of the complete-case estimates was further assessed through cluster-bootstrap resampling by participant (2,000 and 5,000 replicates), with sign consistency defined as the proportion of replicates yielding an estimate in the same direction as the complete-case point estimate. For the subgroup analyses, participants were stratified by educational attainment (low vs. high) and age at the end of follow-up (<65 vs. ≥65 years), and the Callaway and Sant’Anna estimator was applied separately within each stratum. Given the modest and variable stratum sizes, these analyses were regarded as exploratory and hypothesis-generating rather than as formal tests of interaction. All analyses were performed in R version 4.4.2 using *tidyverse* version 2.0.0, *readxl* version 1.4.5, *did* version 2.3.0 and *boot* version 1.3.32.

### 2.6 Ethics

The EPIPorto study protocol was approved by the Ethics Committee of Hospital de São João, Porto, in 1996, and by the Portuguese Data Protection Authority (Comissão Nacional de Proteção de Dados). All participants provided written informed consent at every wave, and the study was conducted in accordance with the Declaration of Helsinki.

## 3. Results

### 3.1 Sample characteristics

After excluding 40 participants who moved to a census block group with a different gentrification classification during the post-exposure period and 47 whose census block group of residence during that period could not be determined, analytical samples ranged from 207 to 620 participants, depending on the availability of complete repeated measurements and covariate data (Table 1). At baseline, 26.2% of participants with a determinable exposure classification lived in gentrifying census block groups.

**Table 1.** Baseline characteristics of participants included in or excluded from the complete-case analysis.

|  | **Alcohol use** (n=2485) |  |  | **Tobacco use** (n=2485) |  |  | **Total cholesterol** (n=2485) |  |  | **HDL cholesterol** (n=2485) |  |  | **LDL cholesterol** (n=2485) |  |  |
| **Classification** | **Included** | **Excluded** | ***p*** | **Included** | **Excluded** | ***p*** | **Included** | **Excluded** | ***p*** | **Included** | **Excluded** | ***p*** | **Included** | **Excluded** | ***p*** |
|---|---|---|---|---|---|---|---|---|---|---|---|---|---|---|---|
| **n** | 607 | 1,878 |  | 620 | 1,865 |  | 214 | 2,271 |  | 209 | 2,276 |  | 207 | 2,278 |  |
| **Age (mean ± SD)** | 47.0 ± 11.2 | 54.8 ± 16.2 | <0.001 | 47.1 ± 11.1 | 54.8 ± 16.2 | <0.001 | 43.5 ± 8.9 | 53.8 ± 15.7 | <0.001 | 43.5 ± 9.0 | 53.8 ± 15.7 | <0.001 | 43.6 ± 9.0 | 53.7 ± 15.7 | <0.001 |
| **Sex, n (%)** |  |  |  |  |  |  |  |  |  |  |  |  |  |  |  |
| Female | 384 (63.3) | 1,157 (61.6) | 0.495 | 391 (63.1) | 1,150 (61.7) | 0.565 | 125 (58.4) | 1,416 (62.4) | 0.288 | 124 (59.3) | 1,417 (62.3) | 0.447 | 123 (59.4) | 1,418 (62.2) | 0.467 |
| Male | 223 (36.7) | 721 (38.4) |  | 229 (36.9) | 715 (38.3) |  | 89 (41.6) | 855 (37.6) |  | 85 (40.7) | 859 (37.7) |  | 84 (40.6) | 860 (37.8) |  |
| **Education, n (%)** |  |  |  |  |  |  |  |  |  |  |  |  |  |  |  |
| Low | 282 (46.5) | 1,233 (65.7) | <0.001 | 287 (46.3) | 1,228 (65.8) | <0.001 | 93 (43.5) | 1,422 (62.6) | <0.001 | 91 (43.5) | 1,424 (62.6) | <0.001 | 90 (43.5) | 1,425 (62.6) | <0.001 |
| Medium | 110 (18.1) | 261 (13.9) |  | 113 (18.2) | 258 (13.8) |  | 46 (21.5) | 325 (14.3) |  | 45 (21.5) | 326 (14.3) |  | 45 (21.7) | 326 (14.3) |  |
| High | 215 (35.4) | 384 (20.4) |  | 220 (35.5) | 379 (20.3) |  | 75 (35.0) | 524 (23.1) |  | 73 (34.9) | 526 (23.1) |  | 72 (34.8) | 527 (23.1) |  |
| **Marital status, n (%)** |  |  |  |  |  |  |  |  |  |  |  |  |  |  |  |
| Without partner | 153 (25.2) | 649 (34.6) | <0.001 | 154 (24.8) | 648 (34.7) | <0.001 | 49 (22.9) | 753 (33.2) | 0.003 | 48 (23.0) | 754 (33.1) | 0.003 | 49 (23.7) | 753 (33.1) | 0.007 |
| With partner | 454 (74.8) | 1,229 (65.4) |  | 466 (75.2) | 1,217 (65.3) |  | 165 (77.1) | 1,518 (66.8) |  | 161 (77.0) | 1,522 (66.9) |  | 158 (76.3) | 1,525 (66.9) |  |
| **Employment, n (%)** |  |  |  |  |  |  |  |  |  |  |  |  |  |  |  |
| Employed | 447 (73.6) | 861 (45.8) | <0.001 | 453 (73.1) | 855 (45.8) | <0.001 | 173 (80.8) | 1,135 (50.0) | <0.001 | 168 (80.4) | 1,140 (50.1) | <0.001 | 165 (79.7) | 1,143 (50.2) | <0.001 |
| Unemployed | 20 (3.3) | 77 (4.1) |  | 22 (3.5) | 75 (4.0) |  | 10 (4.7) | 87 (3.8) |  | 10 (4.8) | 87 (3.8) |  | 11 (5.3) | 86 (3.8) |  |
| Other | 140 (23.1) | 940 (50.1) |  | 145 (23.4) | 935 (50.1) |  | 31 (14.5) | 1,049 (46.2) |  | 31 (14.8) | 1,049 (46.1) |  | 31 (15.0) | 1,049 (46.0) |  |
| **Panel B. Body mass index, overweight/obesity, systolic and diastolic blood pressure, and nightly sleep duration.** |  |  |  |  |  |  |  |  |  |  |  |  |  |  |  |
|  | **Body mass index** (n=2485) |  |  | **Overweight/obesity** (n=2485) |  |  | **Systolic blood pressure** (n=2485) |  |  | **Diastolic blood pressure** (n=2485) |  |  | **Nightly sleep duration** (n=2485) |  |  |
| **Classification** | **Included** | **Excluded** | ***p*** | **Included** | **Excluded** | ***p*** | **Included** | **Excluded** | ***p*** | **Included** | **Excluded** | ***p*** | **Included** | **Excluded** | ***p*** |
| **n** | 587 | 1,898 |  | 587 | 1,898 |  | 545 | 1,940 |  | 544 | 1,941 |  | 599 | 1,886 |  |
| **Age (mean ± SD)** | 46.7 ± 10.9 | 54.8 ± 16.2 | <0.001 | 46.7 ± 10.9 | 54.8 ± 16.2 | <0.001 | 52.9 ± 8.7 | 52.9 ± 16.9 | 0.933 | 53.0 ± 8.7 | 52.9 ± 16.9 | 0.880 | 46.6 ± 10.9 | 54.9 ± 16.2 | <0.001 |
| **Sex, n (%)** |  |  |  |  |  |  |  |  |  |  |  |  |  |  |  |
| Female | 365 (62.2) | 1,176 (62.0) | 0.962 | 365 (62.2) | 1,176 (62.0) | 0.962 | 336 (61.7) | 1,205 (62.1) | 0.884 | 335 (61.6) | 1,206 (62.1) | 0.854 | 379 (63.3) | 1,162 (61.6) | 0.496 |
| Male | 222 (37.8) | 722 (38.0) |  | 222 (37.8) | 722 (38.0) |  | 209 (38.3) | 735 (37.9) |  | 209 (38.4) | 735 (37.9) |  | 220 (36.7) | 724 (38.4) |  |
| **Education, n (%)** |  |  |  |  |  |  |  |  |  |  |  |  |  |  |  |
| Low | 272 (46.3) | 1,243 (65.5) | <0.001 | 272 (46.3) | 1,243 (65.5) | <0.001 | 318 (58.3) | 1,197 (61.7) | 0.351 | 318 (58.5) | 1,197 (61.7) | 0.387 | 276 (46.1) | 1,239 (65.7) | <0.001 |
| Medium | 106 (18.1) | 265 (14.0) |  | 106 (18.1) | 265 (14.0) |  | 85 (15.6) | 286 (14.7) |  | 85 (15.6) | 286 (14.7) |  | 110 (18.4) | 261 (13.8) |  |
| High | 209 (35.6) | 390 (20.5) |  | 209 (35.6) | 390 (20.5) |  | 142 (26.1) | 457 (23.6) |  | 141 (25.9) | 458 (23.6) |  | 213 (35.6) | 386 (20.5) |  |
| **Marital status, n (%)** |  |  |  |  |  |  |  |  |  |  |  |  |  |  |  |
| Without partner | 142 (24.2) | 660 (34.8) | <0.001 | 142 (24.2) | 660 (34.8) | <0.001 | 112 (20.6) | 690 (35.6) | <0.001 | 112 (20.6) | 690 (35.5) | <0.001 | 150 (25.0) | 652 (34.6) | <0.001 |
| With partner | 445 (75.8) | 1,238 (65.2) |  | 445 (75.8) | 1,238 (65.2) |  | 433 (79.4) | 1,250 (64.4) |  | 432 (79.4) | 1,251 (64.5) |  | 449 (75.0) | 1,234 (65.4) |  |
| **Employment, n (%)** |  |  |  |  |  |  |  |  |  |  |  |  |  |  |  |
| Employed | 438 (74.6) | 870 (45.8) | <0.001 | 438 (74.6) | 870 (45.8) | <0.001 | 355 (65.1) | 953 (49.1) | <0.001 | 354 (65.1) | 954 (49.1) | <0.001 | 445 (74.3) | 863 (45.8) | <0.001 |
| Unemployed | 22 (3.7) | 75 (4.0) |  | 22 (3.7) | 75 (4.0) |  | 25 (4.6) | 72 (3.7) |  | 25 (4.6) | 72 (3.7) |  | 21 (3.5) | 76 (4.0) |  |
| Other | 127 (21.6) | 953 (50.2) |  | 127 (21.6) | 953 (50.2) |  | 165 (30.3) | 915 (47.2) |  | 165 (30.3) | 915 (47.1) |  | 133 (22.2) | 947 (50.2) |  |

*"Included" denotes participants in the complete-case analysis of the corresponding outcome; "Excluded" denotes cohort participants without complete data for that outcome. Continuous variables (age) as mean ± SD; categorical variables as n (%). p-values from Welch's t-tests (age) and chi-squared tests (categorical variables). Education: low (≤9 years, ISCED 0–2), medium (10–12 years, ISCED 3–4), high (>12 years, ISCED 5–8); ISCED, International Standard Classification of Education.*

For most outcomes, participants included in the complete-case analyses were significantly younger, had higher educational attainment, and were more likely to be partnered and employed than those excluded because of missing data or loss to follow-up, although age and educational attainment did not differ for blood pressure (Table 1), suggesting selective loss to follow-up and supporting the use of IPCW. Participants living in gentrifying census block groups had lower educational attainment than those living in non-gentrifying ones, whereas age, sex, marital status and employment status were broadly comparable between groups (Table 2). Characteristics by study group at subsequent waves and unadjusted outcome means and proportions by wave and study group are reported in Supplementary Tables S2–S5 and S6, respectively.

**Table 2.**
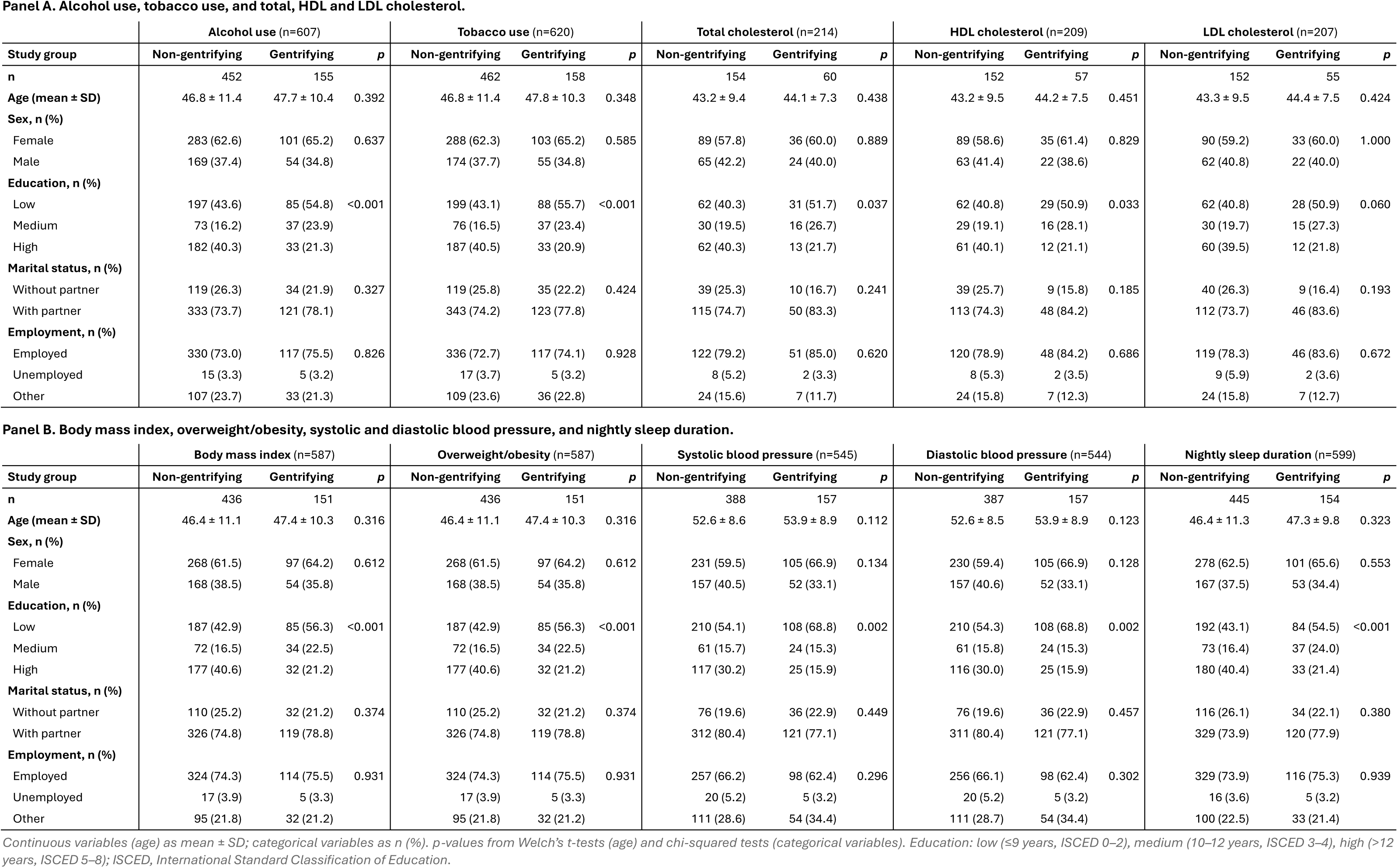
Characteristics by study group within the analytical sample of each health outcome, at baseline.

| Panel A. Alcohol use, tobacco use, and total, HDL and LDL cholesterol. |  |  |  |  |  |  |  |  |  |  |  |  |  |  |  |
|---|---|---|---|---|---|---|---|---|---|---|---|---|---|---|---|
|  | **Alcohol use** (n=607) |  |  | **Tobacco use** (n=620) |  |  | **Total cholesterol** (n=214) |  |  | **HDL cholesterol** (n=209) |  |  | **LDL cholesterol** (n=207) |  |  |
| **Study group** | **Non-gentrifying** | **Gentrifying** | ***p*** | **Non-gentrifying** | **Gentrifying** | ***p*** | **Non-gentrifying** | **Gentrifying** | ***p*** | **Non-gentrifying** | **Gentrifying** | ***p*** | **Non-gentrifying** | **Gentrifying** | ***p*** |
| **n** | 452 | 155 |  | 462 | 158 |  | 154 | 60 |  | 152 | 57 |  | 152 | 55 |  |
| **Age (mean ± SD)** | 46.8 ± 11.4 | 47.7 ± 10.4 | 0.392 | 46.8 ± 11.4 | 47.8 ± 10.3 | 0.348 | 43.2 ± 9.4 | 44.1 ± 7.3 | 0.438 | 43.2 ± 9.5 | 44.2 ± 7.5 | 0.451 | 43.3 ± 9.5 | 44.4 ± 7.5 | 0.424 |
| **Sex, n (%)** |  |  |  |  |  |  |  |  |  |  |  |  |  |  |  |
| Female | 283 (62.6) | 101 (65.2) | 0.637 | 288 (62.3) | 103 (65.2) | 0.585 | 89 (57.8) | 36 (60.0) | 0.889 | 89 (58.6) | 35 (61.4) | 0.829 | 90 (59.2) | 33 (60.0) | 1.000 |
| Male | 169 (37.4) | 54 (34.8) |  | 174 (37.7) | 55 (34.8) |  | 65 (42.2) | 24 (40.0) |  | 63 (41.4) | 22 (38.6) |  | 62 (40.8) | 22 (40.0) |  |
| **Education, n (%)** |  |  |  |  |  |  |  |  |  |  |  |  |  |  |  |
| Low | 197 (43.6) | 85 (54.8) | <0.001 | 199 (43.1) | 88 (55.7) | <0.001 | 62 (40.3) | 31 (51.7) | 0.037 | 62 (40.8) | 29 (50.9) | 0.033 | 62 (40.8) | 28 (50.9) | 0.060 |
| Medium | 73 (16.2) | 37 (23.9) |  | 76 (16.5) | 37 (23.4) |  | 30 (19.5) | 16 (26.7) |  | 29 (19.1) | 16 (28.1) |  | 30 (19.7) | 15 (27.3) |  |
| High | 182 (40.3) | 33 (21.3) |  | 187 (40.5) | 33 (20.9) |  | 62 (40.3) | 13 (21.7) |  | 61 (40.1) | 12 (21.1) |  | 60 (39.5) | 12 (21.8) |  |
| **Marital status, n (%)** |  |  |  |  |  |  |  |  |  |  |  |  |  |  |  |
| Without partner | 119 (26.3) | 34 (21.9) | 0.327 | 119 (25.8) | 35 (22.2) | 0.424 | 39 (25.3) | 10 (16.7) | 0.241 | 39 (25.7) | 9 (15.8) | 0.185 | 40 (26.3) | 9 (16.4) | 0.193 |
| With partner | 333 (73.7) | 121 (78.1) |  | 343 (74.2) | 123 (77.8) |  | 115 (74.7) | 50 (83.3) |  | 113 (74.3) | 48 (84.2) |  | 112 (73.7) | 46 (83.6) |  |
| **Employment, n (%)** |  |  |  |  |  |  |  |  |  |  |  |  |  |  |  |
| Employed | 330 (73.0) | 117 (75.5) | 0.826 | 336 (72.7) | 117 (74.1) | 0.928 | 122 (79.2) | 51 (85.0) | 0.620 | 120 (78.9) | 48 (84.2) | 0.686 | 119 (78.3) | 46 (83.6) | 0.672 |
| Unemployed | 15 (3.3) | 5 (3.2) |  | 17 (3.7) | 5 (3.2) |  | 8 (5.2) | 2 (3.3) |  | 8 (5.3) | 2 (3.5) |  | 9 (5.9) | 2 (3.6) |  |
| Other | 107 (23.7) | 33 (21.3) |  | 109 (23.6) | 36 (22.8) |  | 24 (15.6) | 7 (11.7) |  | 24 (15.8) | 7 (12.3) |  | 24 (15.8) | 7 (12.7) |  |
| **Panel B. Body mass index, overweight/obesity, systolic and diastolic blood pressure, and nightly sleep duration.** |  |  |  |  |  |  |  |  |  |  |  |  |  |  |  |
|  | **Body mass index** (n=587) |  |  | **Overweight/obesity** (n=587) |  |  | **Systolic blood pressure** (n=545) |  |  | **Diastolic blood pressure** (n=544) |  |  | **Nightly sleep duration** (n=599) |  |  |
| **Study group** | **Non-gentrifying** | **Gentrifying** | ***p*** | **Non-gentrifying** | **Gentrifying** | ***p*** | **Non-gentrifying** | **Gentrifying** | ***p*** | **Non-gentrifying** | **Gentrifying** | ***p*** | **Non-gentrifying** | **Gentrifying** | ***p*** |
| **n** | 436 | 151 |  | 436 | 151 |  | 388 | 157 |  | 387 | 157 |  | 445 | 154 |  |
| **Age (mean ± SD)** | 46.4 ± 11.1 | 47.4 ± 10.3 | 0.316 | 46.4 ± 11.1 | 47.4 ± 10.3 | 0.316 | 52.6 ± 8.6 | 53.9 ± 8.9 | 0.112 | 52.6 ± 8.5 | 53.9 ± 8.9 | 0.123 | 46.4 ± 11.3 | 47.3 ± 9.8 | 0.323 |
| **Sex, n (%)** |  |  |  |  |  |  |  |  |  |  |  |  |  |  |  |
| Female | 268 (61.5) | 97 (64.2) | 0.612 | 268 (61.5) | 97 (64.2) | 0.612 | 231 (59.5) | 105 (66.9) | 0.134 | 230 (59.4) | 105 (66.9) | 0.128 | 278 (62.5) | 101 (65.6) | 0.553 |
| Male | 168 (38.5) | 54 (35.8) |  | 168 (38.5) | 54 (35.8) |  | 157 (40.5) | 52 (33.1) |  | 157 (40.6) | 52 (33.1) |  | 167 (37.5) | 53 (34.4) |  |
| **Education, n (%)** |  |  |  |  |  |  |  |  |  |  |  |  |  |  |  |
| Low | 187 (42.9) | 85 (56.3) | <0.001 | 187 (42.9) | 85 (56.3) | <0.001 | 210 (54.1) | 108 (68.8) | 0.002 | 210 (54.3) | 108 (68.8) | 0.002 | 192 (43.1) | 84 (54.5) | <0.001 |
| Medium | 72 (16.5) | 34 (22.5) |  | 72 (16.5) | 34 (22.5) |  | 61 (15.7) | 24 (15.3) |  | 61 (15.8) | 24 (15.3) |  | 73 (16.4) | 37 (24.0) |  |
| High | 177 (40.6) | 32 (21.2) |  | 177 (40.6) | 32 (21.2) |  | 117 (30.2) | 25 (15.9) |  | 116 (30.0) | 25 (15.9) |  | 180 (40.4) | 33 (21.4) |  |
| **Marital status, n (%)** |  |  |  |  |  |  |  |  |  |  |  |  |  |  |  |
| Without partner | 110 (25.2) | 32 (21.2) | 0.374 | 110 (25.2) | 32 (21.2) | 0.374 | 76 (19.6) | 36 (22.9) | 0.449 | 76 (19.6) | 36 (22.9) | 0.457 | 116 (26.1) | 34 (22.1) | 0.380 |
| With partner | 326 (74.8) | 119 (78.8) |  | 326 (74.8) | 119 (78.8) |  | 312 (80.4) | 121 (77.1) |  | 311 (80.4) | 121 (77.1) |  | 329 (73.9) | 120 (77.9) |  |
| **Employment, n (%)** |  |  |  |  |  |  |  |  |  |  |  |  |  |  |  |
| Employed | 324 (74.3) | 114 (75.5) | 0.931 | 324 (74.3) | 114 (75.5) | 0.931 | 257 (66.2) | 98 (62.4) | 0.296 | 256 (66.1) | 98 (62.4) | 0.302 | 329 (73.9) | 116 (75.3) | 0.939 |
| Unemployed | 17 (3.9) | 5 (3.3) |  | 17 (3.9) | 5 (3.3) |  | 20 (5.2) | 5 (3.2) |  | 20 (5.2) | 5 (3.2) |  | 16 (3.6) | 5 (3.2) |  |
| Other | 95 (21.8) | 32 (21.2) |  | 95 (21.8) | 32 (21.2) |  | 111 (28.6) | 54 (34.4) |  | 111 (28.7) | 54 (34.4) |  | 100 (22.5) | 33 (21.4) |  |

*Continuous variables (age) as mean ± SD; categorical variables as n (%). p-values from Welch's t-tests (age) and chi-squared tests (categorical variables). Education: low (≤9 years, ISCED 0–2), medium (10–12 years, ISCED 3–4), high (>12 years, ISCED 5–8); ISCED, International Standard Classification of Education.*

### 3.2 Impact of gentrification on health outcomes

In the primary IPCW analysis, the pre-test detected departures from parallel trends for current tobacco use and LDL cholesterol in the adjusted model and for systolic blood pressure in both models (p<0.05; Supplementary Table S7), whose ATTs were therefore not interpreted causally, but not for the remaining outcomes, including nightly sleep duration. Residence in a gentrifying neighbourhood was associated with a statistically significant reduction in nightly sleep duration (adjusted ATT = −0.346 h; 95% CI −0.650, −0.042), corresponding to about 21 fewer minutes of sleep. No statistically significant associations were observed for the remaining outcomes in either model (Table 3; wave-specific estimates in Supplementary Table S8).

**Table 3.**
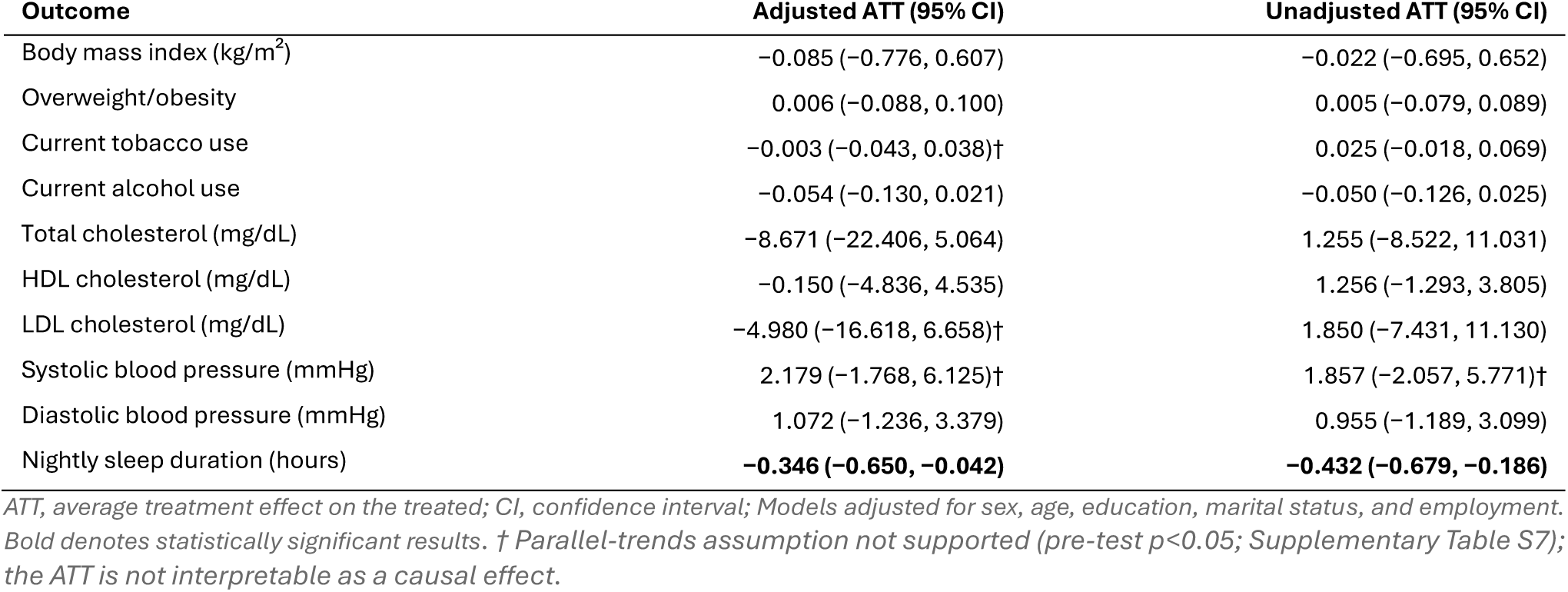
ATT of gentrification on the ten health outcomes in the IPCW analysis (adjusted and unadjusted models).

### 3.3 Impact of gentrification on health outcomes: exploratory subgroup analyses

In the adjusted IPCW model, residence in a gentrifying census block group was associated with shorter nightly sleep duration among participants aged ≥65 years (−0.433 h; 95% CI −0.859, −0.007) and those with lower educational attainment (−0.415 h; 95% CI −0.806, −0.024), but not among participants aged <65 years or those with higher educational attainment (Table 4). Nightly sleep duration was the only outcome with statistically significant estimates within any stratum, and the unadjusted IPCW model showed the same pattern (Supplementary Table S20). The parallel-trends assumption was supported for nightly sleep duration in all four strata but not for a few non-significant estimates of other outcomes, mostly systolic blood pressure among participants aged ≥65 years (Supplementary Tables S18–S19).

**Table 4.** Final aggregate ATT for the IPCW analysis within subgroups (adjusted models).

| **Outcome** | **Lower education** | **Higher education** | **Older (≥65 years)** | **Younger (<65 years)** |
|---|---|---|---|---|
|  | ATT (95% CI) | ATT (95% CI) | ATT (95% CI) | ATT (95% CI) |
| Body mass index (kg/m²) | −0.169 (−1.127, 0.789) | 0.528 (−0.536, 1.591) | −0.447 (−1.440, 0.546) | 0.937 (−0.009, 1.883) |
| Overweight/obesity | 0.012 (−0.113, 0.137) | 0.067 (−0.083, 0.217) | 0.005 (−0.126, 0.137) | 0.033 (−0.084, 0.151) |
| Total cholesterol (mg/dL) | −8.459 (−31.247, 14.329) | 4.766 (−12.761, 22.293) | −8.610 (−33.030, 15.809) | 2.155 (−9.385, 13.696) |
| HDL cholesterol (mg/dL) | 2.981 (−7.392, 13.355) | 0.711 (−4.407, 5.829) | 3.632 (−6.793, 14.056) | 0.483 (−3.418, 4.385) |
| LDL cholesterol (mg/dL) | 0.213 (−19.173, 19.598) | 0.621 (−14.858, 16.100) | −7.569 (−27.294, 12.155) | 4.039 (−5.895, 13.973) |
| Systolic blood pressure (mmHg) | 3.669 (−1.216, 8.554) | 1.215 (−4.892, 7.322) | 2.169 (−2.489, 6.827) | 0.874 (−4.563, 6.311) |
| Diastolic blood pressure (mmHg) | 1.033 (−1.854, 3.919) | 0.642 (−2.718, 4.002) | 1.001 (−1.654, 3.656) | 0.531 (−2.964, 4.026) |
| Current tobacco use | −0.002 (−0.052, 0.047) | 0.113 (−0.004, 0.230) | 0.004 (−0.039, 0.047) | 0.013 (−0.097, 0.124) |
| Current alcohol use | −0.074 (−0.177, 0.030) | 0.048 (−0.078, 0.174) | −0.051 (−0.150, 0.049) | 0.037 (−0.059, 0.133) |
| Nightly sleep duration (hours) | **−0.415 (−0.806, −0.024)** | 0.191 (−0.315, 0.698) | **−0.433 (−0.859, −0.007)** | −0.188 (−0.568, 0.193) |

*Education: lower education (≤9 years, ISCED 0–2), higher education (>12 years, ISCED 5–8); ATT, average treatment effect on the treated; CI, confidence interval; Models adjusted for sex, age, education, marital status, and employment. Bold denotes statistically significant results. Within each stratum, the stratifying variable was omitted from the adjustment set because it was constant.*

### 3.4 Sensitivity and robustness analyses

The results for nightly sleep duration were robust across alternative exposure definitions. Under both the stricter (≥1.5 SD above the mean) and the broader (upper quartile of the index) definitions, nightly sleep duration remained the only outcome significantly associated with gentrification in the adjusted and unadjusted IPCW analyses, with adjusted estimates similar to the primary analysis (−0.349 h; 95% CI −0.658, −0.040 and −0.350 h; 95% CI −0.697, −0.003, respectively), and the parallel-trends assumption was supported for all outcomes (Supplementary Tables S25–S29). The corresponding complete-case estimates for sleep duration were in the same direction but smaller and not statistically significant (−0.129 h; 95% CI −0.445, 0.187 and −0.158 h; 95% CI −0.431, 0.115, respectively; Supplementary Table S27).

Within subgroups, alternative exposure definitions generally reproduced the pattern observed in the primary subgroup analysis, although statistical significance varied across specifications (Supplementary Tables S38–S45). Under the upper-quartile definition, the parallel-trends assumption was supported for nightly sleep duration in all strata (Supplementary Tables S34–S37). The unadjusted IPCW estimates remained statistically significant among participants aged ≥65 years (−0.424 h; 95% CI −0.775, −0.073) and those with lower educational attainment (−0.418 h; 95% CI −0.819, −0.016). The corresponding adjusted estimates were similar in magnitude but no longer statistically significant (−0.382 h; 95% CI −0.820, 0.057 and −0.326 h; 95% CI −0.803, 0.152, respectively).

Under the ≥1.5 SD definition, IPCW estimates among participants aged ≥65 years were larger in both the unadjusted (−0.501 h; 95% CI −0.893, −0.109) and adjusted models (−0.543 h; 95% CI −1.048, −0.038). However, the parallel-trends assumption was violated in this stratum under both models (p=0.013 and p=0.005, respectively; Supplementary Tables S32 and S33). In the corresponding complete-case analyses, estimates among participants aged ≥65 years were statistically significant and the parallel-trends assumption was supported in both the unadjusted (−0.401 h; 95% CI −0.775, −0.028) and adjusted models (−0.373 h; 95% CI −0.685, −0.061) (Supplementary Tables S30, S31, S38 and S39).

Isolated statistically significant estimates for other outcomes were observed only under the alternative exposure definitions. Under the ≥1.5 SD definition, these involved total and HDL cholesterol and systolic blood pressure among participants with lower educational attainment or aged ≥65 years, and total and LDL cholesterol and diastolic blood pressure among participants aged <65 years. Under the upper-quartile definition, they involved systolic blood pressure among participants aged <65 years and diastolic blood pressure among those with higher educational attainment and occurred only in the unadjusted models (Supplementary Tables S38–S42 and S44). None was reproduced in the primary analysis, several were accompanied by violations of the parallel-trends assumption, and their directions were inconsistent across exposure definitions and model specifications. As these subgroup analyses were exploratory, not powered to formally test interaction effects, and not corrected for multiple comparisons, they should be interpreted as hypothesis-generating.

When follow-up wave 2 was re-included as the first post-exposure wave, no statistically significant associations were observed for any outcome in either the IPCW or complete-case analyses, adjusted or unadjusted, and the parallel-trends assumption was supported throughout (Supplementary Tables S21– S24). This analysis was not informative for nightly sleep duration, which was not assessed at wave 2. Retaining the 40 participants who moved between differently classified census block groups did not materially change the results: nightly sleep duration remained the only outcome significantly associated with gentrification (adjusted IPCW ATT = −0.316 h; 95% CI −0.605, −0.027), and the pattern of parallel-trends pre-test results was largely unchanged (Supplementary Table S46).

In the complete-case analyses of the overall effect, no statistically significant associations were observed for any outcome (Supplementary Tables S9–S12). For nightly sleep duration, the adjusted (−0.252 h; 95% CI −0.509, 0.005) and unadjusted (−0.244 h; 95% CI −0.513, 0.025) estimates were in the same direction as in the primary analysis but smaller, and the parallel-trends assumption was not supported in either model (p=0.030 and p=0.032, respectively). Within subgroups, complete-case sleep estimates were in the same direction as the IPCW estimates, of similar magnitude among participants with lower educational attainment (adjusted −0.351 h; 95% CI −0.766, 0.063) but attenuated among those aged ≥65 years (adjusted −0.188 h; 95% CI −0.529, 0.154), and not statistically significant in any subgroup or model (Supplementary Tables S14–S17).

Cluster-bootstrap resampling of the complete-case estimates yielded confidence intervals that included the null for all outcomes (Supplementary Table S13). Sign consistency was highest for nightly sleep duration (0.93 with 2,000 and 0.92 with 5,000 replicates), whereas the direction of most other estimates was unstable, consistent with their limited precision. Overall, the reduction in sleep duration was consistent in direction across analytical approaches and exposure definitions but, at the overall level, reached statistical significance only in the IPCW analyses, whereas the null findings for the remaining outcomes were consistent across all sensitivity analyses.

## 4. Discussion

To our knowledge, this is the first quantitative quasi-experimental study to estimate the health impacts of gentrification in Portugal and one of the few conducted in Europe. In this population-based cohort of adults from Porto, residence in a gentrifying neighbourhood was associated with a statistically significant reduction in nightly sleep duration of approximately 21 minutes, after adjustment for individual-level covariates and accounting for selective loss to follow-up, whereas no statistically significant effects were observed for the remaining outcomes.

The observed reduction in sleep is consistent with conceptual frameworks linking gentrification and neighbourhood disruption to health. Gentrification may affect sleep through several mutually reinforcing pathways, including increased nocturnal noise, the expansion of nightlife and tourism-related activity, the growth of short-term rental accommodation and greater housing insecurity, all of which have been linked to poorer sleep among urban residents (Anguelovski et al., 2021; Cole et al., 2021; Santos, Henriques, et al., 2024). In Porto, qualitative studies conducted within the same cohort have documented nocturnal noise, perceived insecurity and housing-related stress as resident-reported consequences of tourism- and student-driven neighbourhood change (Santos, Silva, et al., 2024; Silva et al., 2023), for which the present findings provide quantitative support.

Quantitative evidence linking neighbourhood change to sleep remains scarce and has generally addressed neighbourhood reinvestment rather than gentrification itself. In two low-income Pittsburgh neighbourhoods, residents living closest to major public and private investments were protected against the decline in sleep duration observed among those living farther away, a difference of 18.3 minutes per night, although the neighbourhood-level comparison was null (Dubowitz et al., 2021). In contrast with our findings, an ecological analysis of US census tracts reported a small decline in the prevalence of insufficient sleep in gentrifying areas, although its aggregate design cannot separate improvement among remaining residents from population replacement, and its threshold-based outcome is not directly comparable with mean sleep duration (Candipan et al., 2023).

Most longitudinal studies have instead focused on proximal features of neighbourhood change, such as socioeconomic disadvantage, residential instability, housing insecurity and environmental noise (Kim et al., 2022). Neighbourhood disadvantage and disorder have been associated with shorter and more fragmented sleep, while housing insecurity has been prospectively associated with shorter sleep duration among low-income adults (Bozick et al., 2021). Against this background, the reduction observed in this study is potentially important at the population level: its magnitude is comparable to that reported by Dubowitz et al. (2021), which those authors considered policy-relevant, and insufficient sleep is associated with cardiovascular morbidity and mortality (Cappuccio et al., 2011), although whether a change of this magnitude translates into measurable cardiometabolic effects over the period examined remains uncertain.

In exploratory subgroup analyses, the effect on sleep was statistically significant among older adults and participants with lower educational attainment, the groups considered a priori to be most vulnerable to neighbourhood change, but not among younger or more highly educated participants. Although these analyses were not powered to formally test interaction effects nor adjusted for multiple comparisons, this pattern is consistent with previous evidence of greater susceptibility to neighbourhood change and environmental stressors in these groups (Ribeiro, Fraga, et al., 2022; Santos, Henriques, et al., 2024) and with the expectation that the burdens of neighbourhood upgrading fall disproportionately on those with fewer resources to adapt or relocate (Cole et al., 2021; Cole et al., 2024).

The absence of statistically significant effects on most cardiometabolic and behavioural outcomes is consistent with the broader international literature, in which beneficial, adverse and null effects of gentrification on cardiometabolic health have been reported without a consistent pattern in the direction or magnitude of associations (Delong, 2023; Schnake-Mahl, Jahn, et al., 2020; Tulier et al., 2019), including modestly improved hypertension control in Los Angeles County (Chen et al., 2023) and no association with incident metabolic syndrome among Hispanic/Latino adults (Pichardo et al., 2023).

Several non-mutually exclusive explanations may account for these findings. First, gentrification may exert opposing effects on the same outcome (Breyer C Voss-Andreae, 2013), which may cancel out at the population level. Second, the relevant pathways may operate over different time horizons: the psychosocial mechanisms most directly engaged by transnational gentrification, such as nocturnal noise, insecurity and housing-related stress, may act on sleep over a relatively shorter period, whereas body mass index, cholesterol and blood pressure are shaped by cumulative life-course exposures and change more gradually (Lynch C Smith, 2005). The post-exposure follow-up may therefore have been sufficient to detect effects on sleep but not on downstream cardiometabolic risk factors, for which sleep may itself represent an upstream determinant.

Third, and most importantly, the precision of the estimates was limited by the small size of the exposed group. Confidence intervals for the cardiometabolic outcomes were wide and compatible with effects in either direction, and cluster-bootstrap resampling showed that the direction of most estimates was unstable. Because IPCW reduces bias arising from differential loss to follow-up but does not increase the effective sample size, the null estimates, particularly those for blood pressure and cholesterol, should be interpreted as inconclusive rather than as evidence of no effect.

The discrepancy between the IPCW and complete-case estimates for sleep duration warrants consideration. Both approaches yielded estimates in the same direction, and the difference in statistical significance stemmed not from greater precision of the IPCW analysis, whose confidence interval was in fact wider, but from a larger point estimate (−0.346 h vs −0.252 h). Participants retained in the complete-case samples were younger, more highly educated and more often employed than those lost to follow-up or with missing data (Table 1), whereas the effect on sleep appeared to concentrate among older and less educated participants. By up-weighting retained participants who resembled those lost to follow-up, IPCW restores the contribution of these under-represented groups, which may explain the larger overall estimate; consistently, the complete-case estimate was most attenuated among participants aged ≥65 years (−0.188 h vs −0.433 h in the IPCW analysis). Both complete-case estimates were further limited by violations of the parallel-trends assumption.

Nevertheless, IPCW relies on the assumption that, conditional on measured baseline covariates, loss to follow-up is unrelated to the outcomes; if attrition depended on unmeasured factors, such as health status or residential displacement, residual selection bias cannot be excluded.

### 4.1 Strengths

This study has several methodological strengths. First, its longitudinal, quasi-experimental design, combined with the Callaway and Sant’Anna estimator, addressed key limitations of previous cross-sectional research. Second, combining IPCW to address selective loss to follow-up with cluster-bootstrap resampling to assess estimate stability provided a more rigorous evaluation than is common in gentrification research.

Third, the estimated reduction in sleep duration remained similar and statistically significant in the IPCW analyses when gentrifying neighbourhoods were defined using other thresholds, reducing concerns about the arbitrariness of the primary cut-off. Fourth, the PCA-based gentrification index, previously validated against objective indicators of neighbourhood change and residents’ perceptions (Alves et al., 2026), integrated multiple dimensions of neighbourhood change within a reproducible and theoretically informed framework. Unlike generic measures of socioeconomic change, it incorporated high-income foreign residents and short-term tourist accommodation, capturing the transnational, tourism-driven character of gentrification particularly salient in Southern European cities such as Porto, which may affect health through mechanisms not captured by a purely socioeconomic index.

Finally, the simultaneous assessment of ten outcomes provided a broad overview of potential health impacts within the same population, and five follow-up waves spanning more than two decades enabled the construction of the pre-exposure trajectories required for valid DiD inference.

### 4.2 Limitations

Several limitations should be acknowledged. First, the operationalisation of gentrification remains heterogeneous across studies, limiting direct cross-study comparability (Bhavsar et al., 2020; Mujahid et al., 2019), although this was partly addressed by combining domains frequently used in previous research and by empirically assessing the structure and construct validity of the index (Alves et al., 2026). Second, outcome selection was constrained by the availability of repeated and comparable measurements, which precluded mental health outcomes such as depression and anxiety, for which associations with gentrification have previously been reported (Anguelovski et al., 2021). This is particularly relevant because our results are compatible with an important role of psychosocial pathways, which could therefore only be assessed indirectly.

Third, the post-exposure follow-up period was shorter for blood pressure and alcohol use than for the remaining outcomes, reducing comparability across estimates and potentially limiting the detection of longer-term changes. Fourth, exposure classification was based on residential address and was therefore subject to the Uncertain Geographic Context Problem (Kwan, 2012), as participants’ daily exposure to gentrification-related changes may extend beyond their residential neighbourhood. Fifth, the few participants who moved between differently classified census block groups were excluded from the primary analysis, although retaining them did not materially change the estimates. More importantly, the estimates do not capture the health consequences of displacement, a central mechanism through which gentrification may affect health (Anguelovski et al., 2021), also documented in Porto (Silva et al., 2025), and may therefore underestimate its overall health impact.

Sixth, the small size of the exposed group limited the ability to distinguish the absence of an effect from insufficient statistical precision. Finally, although the difference-in-differences design accounts for time-invariant differences between study groups and the analyses adjusted for measured individual-level covariates, residual confounding due to unmeasured time-varying individual- or neighbourhood-level factors remains possible.

### 4.3 Conclusions

In this population-based cohort, living in a neighbourhood undergoing transnational gentrification in Porto was associated with a reduction of approximately 21 minutes in nightly sleep duration. This effect was consistent in direction across analytical approaches, robust to alternative exposure definitions, and appeared to be concentrated among older adults and those with lower educational attainment. No effects were detected on body mass index, cholesterol, blood pressure, or tobacco and alcohol use, although limited precision and, for some outcomes, violations of the parallel-trends assumption do not allow such effects to be ruled out. Sleep may thus be an early and sensitive marker of the health impact of gentrification and a plausible pathway through which it may widen health inequities among the most vulnerable residents.

The methodological framework applied in this study provides a reproducible approach for examining urban processes as population health exposures. Replication in cohorts with larger exposed groups and longer post-exposure follow-up is needed to clarify the long-term health consequences of gentrification and to support evidence-informed urban policy.

## Declaration of competing interests

The authors declare no competing interests.

## Funding

This study is integrated in the HUG project, supported by the Foundation for Science and Technology (FCT), I.P., through the projects with references UID/04750/2025 and LA/P/0064/2020 and DOI identifiers https://doi.org/10.54499/UID/04750/2025 and https://doi.org/10.54499/LA/P/0064/2020, and project PTDC/GES-OUT/1662/2020 (https://doi.org/10.54499/PTDC/GES-OUT/1662/2020). Ana Isabel Ribeiro was supported by FCT under the Stimulus of Scientific Employment programme (CEECIND/02386/2018). Henrique Alves was supported by National Funds through the FCT grant 2024.06298.BD.

This research also received support from the Centre of Studies in Geography and Spatial Planning (CEGOT), funded by National Funds through the FCT under the reference UIDB/04084/2025. Further funding was provided by the Social Observatory of “La Caixa” through the project Health Outcomes of Touristification and Gentrification in Portuguese Cities: A Nationwide Survey (HOT-CITIES; FP25-1B113), funded under the FP25-1B Quantitative Surveys in the Social Sciences call.

## CRediT author statement

José António Figueiredo: Conceptualization, Data curation, Methodology, Software, Formal analysis, Investigation, Writing – original draft, Writing – review C editing.

Henrique Alves: Conceptualization, Methodology, Software, Formal analysis, Investigation, Writing – original draft, Writing – review C editing.

Carla Moreira: Methodology, Formal analysis, Supervision, Validation, Writing – review C editing.

Ana Isabel Ribeiro: Conceptualization, Methodology, Funding acquisition, Resources, Project administration, Supervision, Validation, Writing – review C editing.

## Data availability

The EPIPorto data are not publicly available because they contain sensitive health information protected under data protection regulations. Data are available from the corresponding author upon reasonable request, subject to approval by the EPIPorto cohort coordinators and to the applicable ethical and data protection requirements.

## Declaration of generative AI and AI-assisted technologies in the manuscript preparation process

During the preparation of this work, the authors used Claude (Anthropic) to review the manuscript text and to assist with the R code used in the statistical analyses. After using this tool, the authors reviewed and edited the content as needed and take full responsibility for the content of the published article.

## Notes

### Competing Interest Statement

The authors have declared no competing interest.

### Summary of Updates

The authors' affiliations have been updated to ensure that the information up to date.

