## Supplementary Material for "Impact of transnational gentrification on the health of adults from the EPIPorto cohort: a quasi-experimental study"

### Index of tables

### Index of figures

Supplementary Figure S1. Thematic map of the spatial distribution of census block groups classified as gentrifying and non-gentrifying in Porto. ........ 5

### Section A. Principal Component Analysis

The gentrification index was derived by Principal Component Analysis (PCA) of census block group indicators. Full details on the construction and validation of the index are reported elsewhere (Alves et al., 2026). Fifteen candidate indicators, each expressed as its 2011 value and its absolute change between 2011 and 2021 (30 variables), were considered, of which 16 were retained following correlation and redundancy analysis. The chosen principal component most clearly captured the temporal socio-demographic and economic transformations characteristic of gentrification, assigning positive weights (component score coefficients) to change in the proportion of residents with tertiary education (0.173), foreign residents from high-income countries (0.160) and young adult residents (0.217), precisely the drivers of the current gentrification process in Porto.

The chosen principal component also showed a high discriminatory capacity for census block groups with lower socioeconomic status in the initial moment, which are more susceptible to gentrification. For example, it assigned a negative weight (−0.082) to the proportion of residents with tertiary education in the initial moment. By contrast, the first component mainly captured census block groups that were already affluent in 2011, whereas the selected component combined the variables that most specifically reflected the multidimensional processes conceptualised as transnational gentrification (Alves et al., 2026). The chosen PC was therefore retained as the basis of the gentrification index.

**Supplementary Table S1.** Variables included in the gentrification index and respective weights (N = 1,443 census block groups).

| Variable | Index weight |
|---|---|
| Residents with Complete Higher Education (proportion in 2011) % | −0.082 |
| Residents with Complete Higher Education (absolute change 2011–2021) | 0.173 |
| Residents in White-Collar jobs (proportion in 2011) %[1] | −0.061 |
| Residents in White-Collar jobs (absolute change 2011–2021)[1] | 0.156 |
| Housing rents above 650€ (proportion in 2011) % | −0.096 |
| Housing rents above 650€ (absolute change 2011–2021) | 0.092 |
| Residents aged 65 years old or over (proportion in 2011) % | 0.193 |
| Residents aged 65 years old or over (absolute change 2011–2021) | −0.149 |
| Residents aged 20–34 years old (proportion in 2011) % | −0.118 |
| Residents aged 20–34 years old (absolute change 2011–2021) | 0.217 |
| Foreign Residents from high-income countries (proportion in 2011) %[2] | −0.027 |
| Foreign Residents from high-income countries (absolute change 2011–2021)[2] | 0.160 |
| Unemployed Residents (from the age group 25–64 years old) (proportion in 2011) % | 0.192 |
| Unemployed Residents (from the age group 25–64 years old) (absolute change 2011–2021) | −0.164 |
| Short-term rentals (proportion in 2011) % | 0.051 |
| Short-term rentals (absolute change 2011–2021) | 0.172 |

*CPP: Portuguese Classification of Occupations. [1] Residents employed in CPP groups 0–2 (higher-skilled occupations). [2] From the concept of foreign residents from high-income countries, Romania and Bulgaria were excluded because, in 2011, there was still a considerable number of immigrants from these countries who did not reflect the gentrification phenomenon — more so given that their numbers decreased markedly between 2011 and 2021. Weights are the component score coefficients of the selected (second) principal component, used to compute the gentrification index (Alves et al., 2026).*

Census block groups classified as gentrifying were predominantly concentrated in the historical centre, along the central and western waterfront, in the university area in the north, and in the historically disadvantaged eastern parishes, consistent with the spatial pattern of touristification, student-driven change, and rehabilitation documented in literature (Sousa & Barcón, 2021). Supplementary Figure S1 provides an illustrative spatial distribution of census block groups classified as gentrifying and non-gentrifying in Porto.

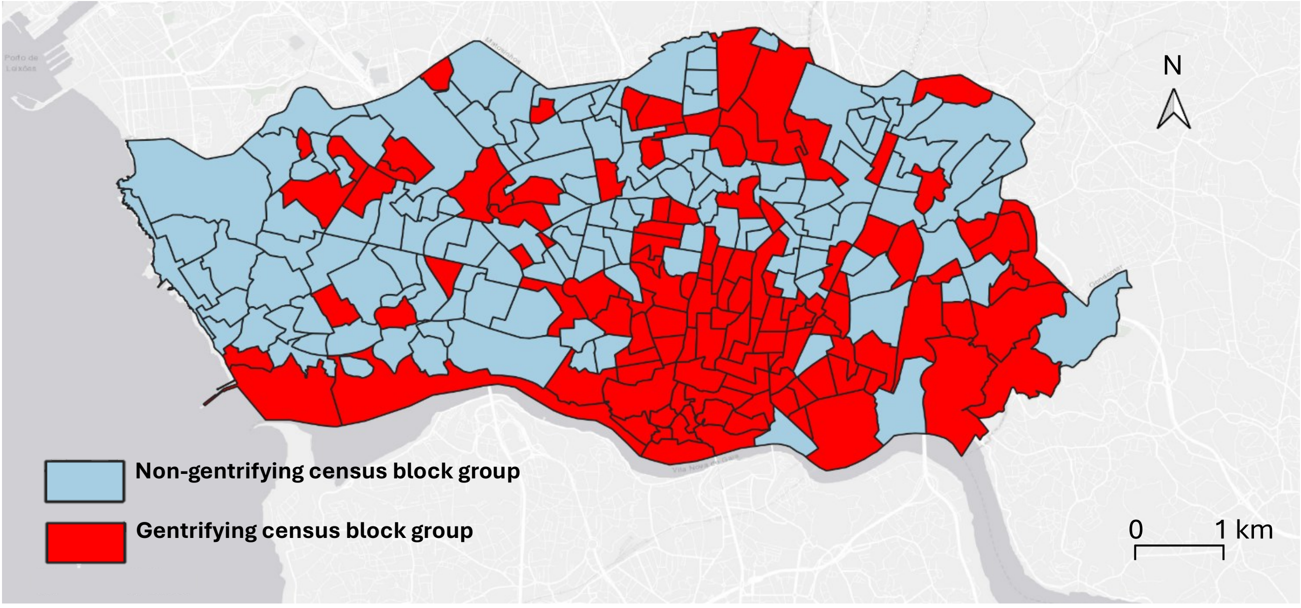

**Supplementary Figure S1.** Thematic map of the spatial distribution of census block groups classified as gentrifying and non-gentrifying in Porto.

### Section B. Definitions and measurement of health outcomes

Ten outcomes with repeated, comparable measurements across waves were analysed. Seven were continuous (body mass index; total, HDL and LDL cholesterol; systolic and diastolic blood pressure; and average nightly sleep duration) and three were binary (overweight/obesity, current tobacco use and current alcohol use).

Body mass index was computed as weight (kg) divided by height squared ($m^2$). Weight was measured with a calibrated TANITA® scale and height standing with a stadiometer. Total and HDL were obtained from serum analyses (mg/dL) by enzymatic colorimetric assay on an Olympus AU5400 analyser, and LDL cholesterol was estimated with the Friedewald formula from total cholesterol, HDL cholesterol and triglycerides. Systolic and diastolic blood pressure (mmHg) were computed as the mean of the repeated measurements available within each wave. Average nightly sleep duration, originally recorded in hours and minutes, was converted to decimal hours.

Overweight/obesity was defined, following the World Health Organization criteria, as body mass index ≥ 25.0 $kg/m^2$ (without further subdivision, to ease interpretation). Current tobacco use and current alcohol use classified participants as active consumers vs non-consumers of, respectively, tobacco and alcoholic beverages at the time of each assessment.

**Supplementary Table S2.** Characteristics by study group within the analytical sample of each health outcome, at follow-up wave 1.

**Panel A. Alcohol use, Tobacco use, Total cholesterol, HDL cholesterol, LDL cholesterol.**

| | **Alcohol use** (n=607) | | | **Tobacco use** (n=620) | | | **Total cholesterol** (n=214) | | | **HDL cholesterol** (n=209) | | | **LDL cholesterol** (n=207) | | |
|---|---|---|---|---|---|---|---|---|---|---|---|---|---|---|---|
| **Study group** | **Non-gentrifying** | **Gentrifying** | ***p*** | **Non-gentrifying** | **Gentrifying** | ***p*** | **Non-gentrifying** | **Gentrifying** | ***p*** | **Non-gentrifying** | **Gentrifying** | ***p*** | **Non-gentrifying** | **Gentrifying** | ***p*** |
| **n** | 452 | 155 | | 462 | 158 | | 154 | 60 | | 152 | 57 | | 152 | 55 | |
| **Age (mean ± SD)** | 51.8 ± 11.9 | 52.3 ± 10.6 | 0.581 | 51.8 ± 11.9 | 52.4 ± 10.5 | 0.520 | 47.8 ± 10.1 | 48.9 ± 7.9 | 0.425 | 47.8 ± 10.1 | 48.6 ± 8.0 | 0.538 | 48.0 ± 10.1 | 48.9 ± 7.9 | 0.499 |
| **Sex, n (%)** | | | | | | | | | | | | | | | |
| Female | 283 (62.6) | 101 (65.2) | 0.637 | 288 (62.3) | 103 (65.2) | 0.585 | 89 (57.8) | 36 (60.0) | 0.889 | 89 (58.6) | 35 (61.4) | 0.829 | 90 (59.2) | 33 (60.0) | 1.000 |
| Male | 169 (37.4) | 54 (34.8) | | 174 (37.7) | 55 (34.8) | | 65 (42.2) | 24 (40.0) | | 63 (41.4) | 22 (38.6) | | 62 (40.8) | 22 (40.0) | |
| **Education, n (%)** | | | | | | | | | | | | | | | |
| Low | 179 (39.6) | 83 (53.5) | <0.001 | 180 (39.0) | 85 (53.8) | <0.001 | 56 (36.4) | 31 (51.7) | 0.022 | 56 (36.8) | 29 (50.9) | 0.023 | 56 (36.8) | 28 (50.9) | 0.041 |
| Medium | 80 (17.7) | 36 (23.2) | | 84 (18.2) | 37 (23.4) | | 31 (20.1) | 15 (25.0) | | 30 (19.7) | 15 (26.3) | | 31 (20.4) | 14 (25.5) | |
| High | 193 (42.7) | 36 (23.2) | | 198 (42.9) | 36 (22.8) | | 67 (43.5) | 14 (23.3) | | 66 (43.4) | 13 (22.8) | | 65 (42.8) | 13 (23.6) | |
| **Marital status, n (%)** | | | | | | | | | | | | | | | |
| Without partner | 124 (27.4) | 41 (26.5) | 0.895 | 124 (26.8) | 42 (26.6) | 1.000 | 38 (24.7) | 12 (20.0) | 0.585 | 38 (25.0) | 11 (19.3) | 0.494 | 39 (25.7) | 10 (18.2) | 0.351 |
| With partner | 328 (72.6) | 114 (73.5) | | 338 (73.2) | 116 (73.4) | | 116 (75.3) | 48 (80.0) | | 114 (75.0) | 46 (80.7) | | 113 (74.3) | 45 (81.8) | |
| **Employment, n (%)** | | | | | | | | | | | | | | | |
| Employed | 280 (61.9) | 99 (63.9) | 0.901 | 286 (61.9) | 98 (62.0) | 0.998 | 115 (74.7) | 44 (73.3) | 0.746 | 114 (75.0) | 41 (71.9) | 0.655 | 114 (75.0) | 39 (70.9) | 0.606 |
| Unemployed | 23 (5.1) | 7 (4.5) | | 23 (5.0) | 8 (5.1) | | 11 (7.1) | 3 (5.0) | | 11 (7.2) | 3 (5.3) | | 11 (7.2) | 3 (5.5) | |
| Other | 149 (33.0) | 49 (31.6) | | 153 (33.1) | 52 (32.9) | | 28 (18.2) | 13 (21.7) | | 27 (17.8) | 13 (22.8) | | 27 (17.8) | 13 (23.6) | |

**Panel B. Body mass index, overweight/obesity, systolic and diastolic blood pressure, and nightly sleep duration.**

| | **Body mass index** (n=587) | | | **Overweight/obesity** (n=587) | | | **Systolic blood pressure** (n=545) | | | **Diastolic blood pressure** (n=544) | | | **Nightly sleep duration** (n=599) | | |
|---|---|---|---|---|---|---|---|---|---|---|---|---|---|---|---|
| **Study group** | **Non-gentrifying** | **Gentrifying** | ***p*** | **Non-gentrifying** | **Gentrifying** | ***p*** | **Non-gentrifying** | **Gentrifying** | ***p*** | **Non-gentrifying** | **Gentrifying** | ***p*** | **Non-gentrifying** | **Gentrifying** | ***p*** |
| **n** | 436 | 151 | | 436 | 151 | | 388 | 157 | | 387 | 157 | | 445 | 154 | |
| **Age (mean ± SD)** | 51.4 ± 11.6 | 52.1 ± 10.5 | 0.482 | 51.4 ± 11.6 | 52.1 ± 10.5 | 0.482 | 57.9 ± 8.6 | 58.8 ± 8.8 | 0.322 | 58.0 ± 8.6 | 58.8 ± 8.8 | 0.345 | 51.3 ± 11.7 | 52.0 ± 10.1 | 0.457 |
| **Sex, n (%)** | | | | | | | | | | | | | | | |
| Female | 268 (61.5) | 97 (64.2) | 0.612 | 268 (61.5) | 97 (64.2) | 0.612 | 231 (59.5) | 105 (66.9) | 0.134 | 230 (59.4) | 105 (66.9) | 0.128 | 278 (62.5) | 101 (65.6) | 0.553 |
| Male | 168 (38.5) | 54 (35.8) | | 168 (38.5) | 54 (35.8) | | 157 (40.5) | 52 (33.1) | | 157 (40.6) | 52 (33.1) | | 167 (37.5) | 53 (34.4) | |
| **Education, n (%)** | | | | | | | | | | | | | | | |
| Low | 168 (38.5) | 82 (54.3) | <0.001 | 168 (38.5) | 82 (54.3) | <0.001 | 188 (48.5) | 103 (65.6) | 0.001 | 188 (48.6) | 103 (65.6) | 0.001 | 174 (39.1) | 81 (52.6) | <0.001 |
| Medium | 80 (18.3) | 36 (23.8) | | 80 (18.3) | 36 (23.8) | | 74 (19.1) | 26 (16.6) | | 74 (19.1) | 26 (16.6) | | 80 (18.0) | 37 (24.0) | |
| High | 188 (43.1) | 33 (21.9) | | 188 (43.1) | 33 (21.9) | | 126 (32.5) | 28 (17.8) | | 125 (32.3) | 28 (17.8) | | 191 (42.9) | 36 (23.4) | |
| **Marital status, n (%)** | | | | | | | | | | | | | | | |
| Without partner | 115 (26.4) | 38 (25.2) | 0.854 | 115 (26.4) | 38 (25.2) | 0.854 | 89 (22.9) | 41 (26.1) | 0.498 | 89 (23.0) | 41 (26.1) | 0.508 | 119 (26.7) | 41 (26.6) | 1.000 |
| With partner | 321 (73.6) | 113 (74.8) | | 321 (73.6) | 113 (74.8) | | 299 (77.1) | 116 (73.9) | | 298 (77.0) | 116 (73.9) | | 326 (73.3) | 113 (73.4) | |
| **Employment, n (%)** | | | | | | | | | | | | | | | |
| Employed | 279 (64.0) | 96 (63.6) | 0.936 | 279 (64.0) | 96 (63.6) | 0.936 | 191 (49.2) | 67 (42.7) | 0.333 | 190 (49.1) | 67 (42.7) | 0.346 | 282 (63.4) | 97 (63.0) | 0.991 |
| Unemployed | 23 (5.3) | 7 (4.6) | | 23 (5.3) | 7 (4.6) | | 19 (4.9) | 7 (4.5) | | 19 (4.9) | 7 (4.5) | | 22 (4.9) | 8 (5.2) | |
| Other | 134 (30.7) | 48 (31.8) | | 134 (30.7) | 48 (31.8) | | 178 (45.9) | 83 (52.9) | | 178 (46.0) | 83 (52.9) | | 141 (31.7) | 49 (31.8) | |

*Continuous variables (age) as mean ± SD; categorical variables as n (%), with percentages within each study group. p-values from Welch's t-tests (age) and chi-squared tests (categorical variables). Education: low (≤9 years, ISCED 0–2), medium (10–12 years, ISCED 3–4), high (>12 years, ISCED 5–8).*

**Supplementary Table S3.** Characteristics by study group within the analytical sample of each health outcome, at follow-up wave 2.

**Panel A. Alcohol use, Tobacco use, Total cholesterol, HDL cholesterol, LDL cholesterol.**

| | **Alcohol use** (n=607) | | | **Tobacco use** (n=620) | | | **Total cholesterol** (n=214) | | | **HDL cholesterol** (n=209) | | | **LDL cholesterol** (n=207) | | |
|---|---|---|---|---|---|---|---|---|---|---|---|---|---|---|---|
| **Study group** | **Non-gentrifying** | **Gentrifying** | ***p*** | **Non-gentrifying** | **Gentrifying** | ***p*** | **Non-gentrifying** | **Gentrifying** | ***p*** | **Non-gentrifying** | **Gentrifying** | ***p*** | **Non-gentrifying** | **Gentrifying** | ***p*** |
| **n** | 452 | 155 | | 462 | 158 | | 154 | 60 | | 152 | 57 | | 152 | 55 | |
| **Age (mean ± SD)** | 60.1 ± 11.5 | 60.1 ± 10.5 | 0.990 | 60.1 ± 11.5 | 60.3 ± 10.3 | 0.867 | 55.9 ± 10.1 | 57.0 ± 7.9 | 0.442 | 55.9 ± 10.1 | 56.7 ± 8.1 | 0.564 | 56.1 ± 10.1 | 57.0 ± 8.0 | 0.545 |
| **Sex, n (%)** | | | 0.637 | | | 0.585 | | | 0.889 | | | 0.829 | | | 1.000 |
| Female | 283 (62.6) | 101 (65.2) | | 288 (62.3) | 103 (65.2) | | 89 (57.8) | 36 (60.0) | | 89 (58.6) | 35 (61.4) | | 90 (59.2) | 33 (60.0) | |
| Male | 169 (37.4) | 54 (34.8) | | 174 (37.7) | 55 (34.8) | | 65 (42.2) | 24 (40.0) | | 63 (41.4) | 22 (38.6) | | 62 (40.8) | 22 (40.0) | |
| **Education, n (%)** | | | <0.001 | | | <0.001 | | | 0.004 | | | 0.012 | | | 0.022 |
| Low | 180 (39.8) | 85 (54.8) | | 182 (39.4) | 88 (55.7) | | 57 (37.0) | 32 (53.3) | | 57 (37.5) | 31 (54.4) | | 57 (37.5) | 30 (54.5) | |
| Medium | 69 (15.3) | 30 (19.4) | | 72 (15.6) | 30 (19.0) | | 23 (14.9) | 14 (23.3) | | 23 (15.1) | 12 (21.1) | | 24 (15.8) | 11 (20.0) | |
| High | 203 (44.9) | 40 (25.8) | | 208 (45.0) | 40 (25.3) | | 74 (48.1) | 14 (23.3) | | 72 (47.4) | 14 (24.6) | | 71 (46.7) | 14 (25.5) | |
| **Marital status, n (%)** | | | 0.546 | | | 0.867 | | | 1.000 | | | 1.000 | | | 0.941 |
| Without partner | 127 (28.1) | 39 (25.2) | | 125 (27.1) | 41 (25.9) | | 32 (20.8) | 13 (21.7) | | 32 (21.1) | 12 (21.1) | | 33 (21.7) | 11 (20.0) | |
| With partner | 325 (71.9) | 116 (74.8) | | 337 (72.9) | 117 (74.1) | | 122 (79.2) | 47 (78.3) | | 120 (78.9) | 45 (78.9) | | 119 (78.3) | 44 (80.0) | |
| **Employment, n (%)** | | | 0.343 | | | 0.234 | | | 0.230 | | | 0.304 | | | 0.386 |
| Employed | 198 (43.8) | 62 (40.0) | | 202 (43.7) | 63 (39.9) | | 92 (59.7) | 31 (51.7) | | 91 (59.9) | 30 (52.6) | | 90 (59.2) | 29 (52.7) | |
| Unemployed | 37 (8.2) | 9 (5.8) | | 37 (8.0) | 8 (5.1) | | 16 (10.4) | 4 (6.7) | | 16 (10.5) | 4 (7.0) | | 16 (10.5) | 4 (7.3) | |
| Other | 217 (48.0) | 84 (54.2) | | 223 (48.3) | 87 (55.1) | | 46 (29.9) | 25 (41.7) | | 45 (29.6) | 23 (40.4) | | 46 (30.3) | 22 (40.0) | |

**Panel B. Body mass index, overweight/obesity, systolic and diastolic blood pressure, and nightly sleep duration.**

| | **Body mass index** (n=587) | | | **Overweight/obesity** (n=587) | | | **Systolic blood pressure** (n=545) | | | **Diastolic blood pressure** (n=544) | | | **Nightly sleep duration** (n=599) | | |
|---|---|---|---|---|---|---|---|---|---|---|---|---|---|---|---|
| **Study group** | **Non-gentrifying** | **Gentrifying** | ***p*** | **Non-gentrifying** | **Gentrifying** | ***p*** | **Non-gentrifying** | **Gentrifying** | ***p*** | **Non-gentrifying** | **Gentrifying** | ***p*** | **Non-gentrifying** | **Gentrifying** | ***p*** |
| **n** | 436 | 151 | | 436 | 151 | | 388 | 157 | | 387 | 157 | | 445 | 154 | |
| **Age (mean ± SD)** | 60.0 ± 11.3 | 60.1 ± 10.3 | 0.948 | 60.0 ± 11.3 | 60.1 ± 10.3 | 0.948 | 66.2 ± 8.4 | 66.5 ± 8.5 | 0.729 | 66.3 ± 8.4 | 66.5 ± 8.5 | 0.766 | 59.8 ± 11.5 | 60.0 ± 10.0 | 0.860 |
| **Sex, n (%)** | | | 0.612 | | | 0.612 | | | 0.134 | | | 0.128 | | | 0.553 |
| Female | 268 (61.5) | 97 (64.2) | | 268 (61.5) | 97 (64.2) | | 231 (59.5) | 105 (66.9) | | 230 (59.4) | 105 (66.9) | | 278 (62.5) | 101 (65.6) | |
| Male | 168 (38.5) | 54 (35.8) | | 168 (38.5) | 54 (35.8) | | 157 (40.5) | 52 (33.1) | | 157 (40.6) | 52 (33.1) | | 167 (37.5) | 53 (34.4) | |
| **Education, n (%)** | | | <0.001 | | | <0.001 | | | <0.001 | | | <0.001 | | | <0.001 |
| Low | 170 (39.0) | 86 (57.0) | | 170 (39.0) | 86 (57.0) | | 194 (50.0) | 102 (65.0) | | 194 (50.1) | 102 (65.0) | | 175 (39.3) | 84 (54.5) | |
| Medium | 68 (15.6) | 28 (18.5) | | 68 (15.6) | 28 (18.5) | | 60 (15.5) | 27 (17.2) | | 60 (15.5) | 27 (17.2) | | 70 (15.7) | 30 (19.5) | |
| High | 198 (45.4) | 37 (24.5) | | 198 (45.4) | 37 (24.5) | | 134 (34.5) | 28 (17.8) | | 133 (34.4) | 28 (17.8) | | 200 (44.9) | 40 (26.0) | |
| **Marital status, n (%)** | | | 0.689 | | | 0.689 | | | 0.905 | | | 0.892 | | | 0.936 |
| Without partner | 116 (26.6) | 37 (24.5) | | 116 (26.6) | 37 (24.5) | | 105 (27.1) | 41 (26.1) | | 105 (27.1) | 41 (26.1) | | 119 (26.7) | 40 (26.0) | |
| With partner | 320 (73.4) | 114 (75.5) | | 320 (73.4) | 114 (75.5) | | 283 (72.9) | 116 (73.9) | | 282 (72.9) | 116 (73.9) | | 326 (73.3) | 114 (74.0) | |
| **Employment, n (%)** | | | 0.194 | | | 0.194 | | | 0.128 | | | 0.134 | | | 0.240 |
| Employed | 197 (45.2) | 61 (40.4) | | 197 (45.2) | 61 (40.4) | | 102 (26.3) | 34 (21.7) | | 101 (26.1) | 34 (21.7) | | 200 (44.9) | 63 (40.9) | |
| Unemployed | 36 (8.3) | 8 (5.3) | | 36 (8.3) | 8 (5.3) | | 25 (6.4) | 5 (3.2) | | 25 (6.5) | 5 (3.2) | | 36 (8.1) | 8 (5.2) | |
| Other | 203 (46.6) | 82 (54.3) | | 203 (46.6) | 82 (54.3) | | 261 (67.3) | 118 (75.2) | | 261 (67.4) | 118 (75.2) | | 209 (47.0) | 83 (53.9) | |

*Continuous variables (age) as mean ± SD; categorical variables as n (%), with percentages within each study group. p-values from Welch's t-tests (age) and chi-squared tests (categorical variables). Education: low (≤9 years, ISCED 0–2), medium (10–12 years, ISCED 3–4), high (>12 years, ISCED 5–8).*

**Supplementary Table S4.** Characteristics by study group within the analytical sample of each health outcome, at follow-up wave 3.

**Panel A. Alcohol use, Tobacco use, Total cholesterol, HDL cholesterol, LDL cholesterol.**

| | **Alcohol use** (n=607) | | | **Tobacco use** (n=620) | | | **Total cholesterol** (n=214) | | | **HDL cholesterol** (n=209) | | | **LDL cholesterol** (n=207) | | |
|---|---|---|---|---|---|---|---|---|---|---|---|---|---|---|---|
| **Study group** | **Non-gentrifying** | **Gentrifying** | ***p*** | **Non-gentrifying** | **Gentrifying** | ***p*** | **Non-gentrifying** | **Gentrifying** | ***p*** | **Non-gentrifying** | **Gentrifying** | ***p*** | **Non-gentrifying** | **Gentrifying** | ***p*** |
| **n** | 452 | 155 | | 462 | 158 | | 154 | 60 | | 152 | 57 | | 152 | 55 | |
| **Age (mean ± SD)** | 63.4 ± 11.8 | 63.9 ± 10.5 | 0.678 | 63.5 ± 11.8 | 64.0 ± 10.3 | 0.593 | 59.6 ± 9.9 | 60.6 ± 7.7 | 0.413 | 59.5 ± 9.9 | 60.4 ± 7.8 | 0.535 | 59.7 ± 10.0 | 60.6 ± 7.7 | 0.496 |
| **Sex, n (%)** | | | | | | | | | | | | | | | |
| Female | 283 (62.6) | 101 (65.2) | 0.637 | 288 (62.3) | 103 (65.2) | 0.585 | 89 (57.8) | 36 (60.0) | 0.889 | 89 (58.6) | 35 (61.4) | 0.829 | 90 (59.2) | 33 (60.0) | 1.000 |
| Male | 169 (37.4) | 54 (34.8) | | 174 (37.7) | 55 (34.8) | | 65 (42.2) | 24 (40.0) | | 63 (41.4) | 22 (38.6) | | 62 (40.8) | 22 (40.0) | |
| **Education, n (%)** | | | | | | | | | | | | | | | |
| Low | 180 (39.8) | 85 (54.8) | <0.001 | 182 (39.4) | 88 (55.7) | <0.001 | 57 (37.0) | 32 (53.3) | 0.004 | 57 (37.5) | 31 (54.4) | 0.012 | 57 (37.5) | 30 (54.5) | 0.022 |
| Medium | 69 (15.3) | 30 (19.4) | | 72 (15.6) | 30 (19.0) | | 23 (14.9) | 14 (23.3) | | 23 (15.1) | 12 (21.1) | | 24 (15.8) | 11 (20.0) | |
| High | 203 (44.9) | 40 (25.8) | | 208 (45.0) | 40 (25.3) | | 74 (48.1) | 14 (23.3) | | 72 (47.4) | 14 (24.6) | | 71 (46.7) | 14 (25.5) | |
| **Marital status, n (%)** | | | | | | | | | | | | | | | |
| Without partner | 144 (31.9) | 48 (31.0) | 0.916 | 144 (31.2) | 49 (31.0) | 1.000 | 40 (26.0) | 14 (23.3) | 0.823 | 40 (26.3) | 13 (22.8) | 0.733 | 41 (27.0) | 12 (21.8) | 0.568 |
| With partner | 308 (68.1) | 107 (69.0) | | 318 (68.8) | 109 (69.0) | | 114 (74.0) | 46 (76.7) | | 112 (73.7) | 44 (77.2) | | 111 (73.0) | 43 (78.2) | |
| **Employment, n (%)** | | | | | | | | | | | | | | | |
| Employed | 191 (42.3) | 56 (36.1) | 0.405 | 194 (42.0) | 55 (34.8) | 0.282 | 87 (56.5) | 30 (50.0) | 0.639 | 86 (56.6) | 29 (50.9) | 0.729 | 85 (55.9) | 28 (50.9) | 0.815 |
| Unemployed | 22 (4.9) | 8 (5.2) | | 24 (5.2) | 9 (5.7) | | 11 (7.1) | 4 (6.7) | | 11 (7.2) | 4 (7.0) | | 10 (6.6) | 4 (7.3) | |
| Other | 239 (52.9) | 91 (58.7) | | 244 (52.8) | 94 (59.5) | | 56 (36.4) | 26 (43.3) | | 55 (36.2) | 24 (42.1) | | 57 (37.5) | 23 (41.8) | |

**Panel B. Body mass index, overweight/obesity, systolic and diastolic blood pressure, and nightly sleep duration.**

| | **Body mass index** (n=587) | | | **Overweight/obesity** (n=587) | | | **Systolic blood pressure** (n=545) | | | **Diastolic blood pressure** (n=544) | | | **Nightly sleep duration (n=599)** | | |
|---|---|---|---|---|---|---|---|---|---|---|---|---|---|---|---|
| **Study group** | **Non-gentrifying** | **Gentrifying** | ***p*** | **Non-gentrifying** | **Gentrifying** | ***p*** | **Non-gentrifying** | **Gentrifying** | ***p*** | **Non-gentrifying** | **Gentrifying** | ***p*** | **Non-gentrifying** | **Gentrifying** | ***p*** |
| **n** | 436 | 151 | | 436 | 151 | | 388 | 157 | | 387 | 157 | | 445 | 154 | |
| **Age (mean ± SD)** | 63.1 ± 11.5 | 63.6 ± 10.3 | 0.558 | 63.1 ± 11.5 | 63.6 ± 10.3 | 0.558 | 69.7 ± 8.6 | 70.3 ± 8.7 | 0.428 | 69.7 ± 8.5 | 70.3 ± 8.7 | 0.455 | 63.0 ± 11.6 | 63.6 ± 9.9 | 0.542 |
| **Sex, n (%)** | | | | | | | | | | | | | | | |
| Female | 268 (61.5) | 97 (64.2) | 0.612 | 268 (61.5) | 97 (64.2) | 0.612 | 231 (59.5) | 105 (66.9) | 0.134 | 230 (59.4) | 105 (66.9) | 0.128 | 278 (62.5) | 101 (65.6) | 0.553 |
| Male | 168 (38.5) | 54 (35.8) | | 168 (38.5) | 54 (35.8) | | 157 (40.5) | 52 (33.1) | | 157 (40.6) | 52 (33.1) | | 167 (37.5) | 53 (34.4) | |
| **Education, n (%)** | | | | | | | | | | | | | | | |
| Low | 170 (39.0) | 86 (57.0) | <0.001 | 170 (39.0) | 86 (57.0) | <0.001 | 194 (50.0) | 102 (65.0) | <0.001 | 194 (50.1) | 102 (65.0) | 0.001 | 175 (39.3) | 84 (54.5) | <0.001 |
| Medium | 68 (15.6) | 28 (18.5) | | 68 (15.6) | 28 (18.5) | | 60 (15.5) | 27 (17.2) | | 60 (15.5) | 27 (17.2) | | 70 (15.7) | 30 (19.5) | |
| High | 198 (45.4) | 37 (24.5) | | 198 (45.4) | 37 (24.5) | | 134 (34.5) | 28 (17.8) | | 133 (34.4) | 28 (17.8) | | 200 (44.9) | 40 (26.0) | |
| **Marital status, n (%)** | | | | | | | | | | | | | | | |
| Without partner | 135 (31.0) | 45 (29.8) | 0.869 | 135 (31.0) | 45 (29.8) | 0.869 | 125 (32.2) | 53 (33.8) | 0.805 | 125 (32.3) | 53 (33.8) | 0.820 | 138 (31.0) | 47 (30.5) | 0.990 |
| With partner | 301 (69.0) | 106 (70.2) | | 301 (69.0) | 106 (70.2) | | 263 (67.8) | 104 (66.2) | | 262 (67.7) | 104 (66.2) | | 307 (69.0) | 107 (69.5) | |
| **Employment, n (%)** | | | | | | | | | | | | | | | |
| Employed | 189 (43.3) | 54 (35.8) | 0.264 | 189 (43.3) | 54 (35.8) | 0.264 | 91 (23.5) | 29 (18.5) | 0.426 | 90 (23.3) | 29 (18.5) | 0.452 | 192 (43.1) | 55 (35.7) | 0.270 |
| Unemployed | 23 (5.3) | 9 (6.0) | | 23 (5.3) | 9 (6.0) | | 11 (2.8) | 4 (2.5) | | 11 (2.8) | 4 (2.5) | | 24 (5.4) | 9 (5.8) | |
| Other | 224 (51.4) | 88 (58.3) | | 224 (51.4) | 88 (58.3) | | 286 (73.7) | 124 (79.0) | | 286 (73.9) | 124 (79.0) | | 229 (51.5) | 90 (58.4) | |

*Continuous variables (age) as mean ± SD; categorical variables as n (%), with percentages within each study group. p-values from Welch's t-tests (age) and chi-squared tests (categorical variables). Education: low (≤9 years, ISCED 0–2), medium (10–12 years, ISCED 3–4), high (>12 years, ISCED 5–8).*

**Supplementary Table S5.** Characteristics by study group within the analytical sample of each health outcome, at follow-up wave 5.

**Panel A. Tobacco use, Total cholesterol, HDL cholesterol, LDL cholesterol, and body mass index.**

| | **Tobacco use** (n=620) | | | **Total cholesterol** (n=214) | | | **HDL cholesterol** (n=209) | | | **LDL cholesterol** (n=207) | | | **Body mass index** (n=587) | | |
|---|---|---|---|---|---|---|---|---|---|---|---|---|---|---|---|
| **Study group** | **Non-gentrifying** | **Gentrifying** | ***p*** | **Non-gentrifying** | **Gentrifying** | ***p*** | **Non-gentrifying** | **Gentrifying** | ***p*** | **Non-gentrifying** | **Gentrifying** | ***p*** | **Non-gentrifying** | **Gentrifying** | ***p*** |
| **n** | 462 | 158 | | 154 | 60 | | 152 | 57 | | 152 | 55 | | 436 | 151 | |
| **Age (mean ± SD)** | 67.9 ± 12.2 | 68.4 ± 10.7 | 0.639 | 63.8 ± 10.3 | 64.8 ± 8.0 | 0.426 | 63.7 ± 10.3 | 64.5 ± 8.2 | 0.550 | 63.9 ± 10.4 | 64.8 ± 8.1 | 0.515 | 67.5 ± 11.9 | 68.0 ± 10.7 | 0.597 |
| **Sex, n (%)** | | | | | | | | | | | | | | | |
| Female | 288 (62.3) | 103 (65.2) | 0.585 | 89 (57.8) | 36 (60.0) | 0.889 | 89 (58.6) | 35 (61.4) | 0.829 | 90 (59.2) | 33 (60.0) | 1.000 | 268 (61.5) | 97 (64.2) | 0.612 |
| Male | 174 (37.7) | 55 (34.8) | | 65 (42.2) | 24 (40.0) | | 63 (41.4) | 22 (38.6) | | 62 (40.8) | 22 (40.0) | | 168 (38.5) | 54 (35.8) | |
| **Education, n (%)** | | | | | | | | | | | | | | | |
| Low | 170 (36.8) | 84 (53.2) | <0.001 | 55 (35.7) | 32 (53.3) | 0.009 | 55 (36.2) | 31 (54.4) | 0.019 | 55 (36.2) | 30 (54.5) | 0.030 | 158 (36.2) | 82 (54.3) | <0.001 |
| Medium | 82 (17.7) | 32 (20.3) | | 25 (16.2) | 13 (21.7) | | 25 (16.4) | 11 (19.3) | | 26 (17.1) | 10 (18.2) | | 78 (17.9) | 30 (19.9) | |
| High | 210 (45.5) | 42 (26.6) | | 74 (48.1) | 15 (25.0) | | 72 (47.4) | 15 (26.3) | | 71 (46.7) | 15 (27.3) | | 200 (45.9) | 39 (25.8) | |
| **Marital status, n (%)** | | | | | | | | | | | | | | | |
| Without partner | 147 (31.8) | 49 (31.0) | 0.929 | 39 (25.3) | 15 (25.0) | 1.000 | 39 (25.7) | 14 (24.6) | 1.000 | 39 (25.7) | 14 (25.5) | 1.000 | 136 (31.2) | 44 (29.1) | 0.712 |
| With partner | 315 (68.2) | 109 (69.0) | | 115 (74.7) | 45 (75.0) | | 113 (74.3) | 43 (75.4) | | 113 (74.3) | 41 (74.5) | | 300 (68.8) | 107 (70.9) | |
| **Employment, n (%)** | | | | | | | | | | | | | | | |
| Employed | 146 (31.6) | 43 (27.2) | 0.562 | 57 (37.0) | 23 (38.3) | 0.429 | 56 (36.8) | 23 (40.4) | 0.440 | 55 (36.2) | 22 (40.0) | 0.532 | 140 (32.1) | 42 (27.8) | 0.596 |
| Unemployed | 17 (3.7) | 7 (4.4) | | 9 (5.8) | 1 (1.7) | | 9 (5.9) | 1 (1.8) | | 8 (5.3) | 1 (1.8) | | 17 (3.9) | 7 (4.6) | |
| Other | 299 (64.7) | 108 (68.4) | | 88 (57.1) | 36 (60.0) | | 87 (57.2) | 33 (57.9) | | 89 (58.6) | 32 (58.2) | | 279 (64.0) | 102 (67.5) | |

**Panel B. Overweight/obesity, and nightly sleep duration.**

| | **Overweight/obesity** (n=587) | | | **Nightly sleep duration** (n=599) | | |
|---|---|---|---|---|---|---|
| **Study group** | **Non-gentrifying** | **Gentrifying** | ***p*** | **Non-gentrifying** | **Gentrifying** | ***p*** |
| **n** | 436 | 151 | | 445 | 154 | |
| **Age (mean ± SD)** | 67.5 ± 11.9 | 68.0 ± 10.7 | 0.597 | 67.4 ± 12.1 | 67.9 ± 10.3 | 0.587 |
| **Sex, n (%)** | | | | | | |
| Female | 268 (61.5) | 97 (64.2) | 0.612 | 278 (62.5) | 101 (65.6) | 0.553 |
| Male | 168 (38.5) | 54 (35.8) | | 167 (37.5) | 53 (34.4) | |
| **Education, n (%)** | | | | | | |
| Low | 158 (36.2) | 82 (54.3) | <0.001 | 163 (36.6) | 80 (51.9) | <0.001 |
| Medium | 78 (17.9) | 30 (19.9) | | 80 (18.0) | 32 (20.8) | |
| High | 200 (45.9) | 39 (25.8) | | 202 (45.4) | 42 (27.3) | |
| **Marital status, n (%)** | | | | | | |
| Without partner | 136 (31.2) | 44 (29.1) | 0.712 | 141 (31.7) | 46 (29.9) | 0.750 |
| With partner | 300 (68.8) | 107 (70.9) | | 304 (68.3) | 108 (70.1) | |
| **Employment, n (%)** | | | | | | |
| Employed | 140 (32.1) | 42 (27.8) | 0.596 | 145 (32.6) | 43 (27.9) | 0.544 |
| Unemployed | 17 (3.9) | 7 (4.6) | | 17 (3.8) | 7 (4.5) | |
| Other | 279 (64.0) | 102 (67.5) | | 283 (63.6) | 104 (67.5) | |

*Continuous variables (age) as mean ± SD; categorical variables as n (%), with percentages within each study group. p-values from Welch's t-tests (age) and chi-squared tests (categorical variables). Education: low (≤9 years, ISCED 0–2), medium (10–12 years, ISCED 3–4), high (>12 years, ISCED 5–8).*

**Supplementary Table S6.** Unadjusted means or proportions of each outcome by wave and study group.

| Outcome / wave | Non-gentrifying | Gentrifying | p-value |
|---|---|---|---|
| **Body mass index (kg/m²), mean (SD)** | | | |
| Baseline | 26.1 (4.3) | 27.6 (5.2) | 0.003 |
| Wave 1 | 26.4 (4.3) | 28.1 (5.7) | 0.001 |
| Wave 3 | 26.9 (4.5) | 28.6 (5.3) | <0.001 |
| Wave 5 | 26.3 (4.3) | 27.4 (4.7) | 0.013 |
| **Overweight/obesity, n (%)** | | | |
| Baseline | 248 (56.9) | 104 (68.9) | 0.010 |
| Wave 1 | 261 (59.9) | 104 (68.9) | 0.049 |
| Wave 3 | 288 (66.1) | 114 (75.5) | 0.031 |
| Wave 5 | 265 (60.8) | 102 (67.5) | 0.139 |
| **Total cholesterol (mg/dL), mean (SD)** | | | |
| Baseline | 214.4 (47.2) | 215.3 (37.3) | 0.881 |
| Wave 1 | 213.6 (38.2) | 223.5 (39.5) | 0.101 |
| Wave 3 | 204.6 (38.1) | 212.8 (46.1) | 0.222 |
| Wave 5 | 189.0 (38.6) | 199.7 (45.3) | 0.108 |
| **HDL cholesterol (mg/dL), mean (SD)** | | | |
| Baseline | 57.7 (16.9) | 56.7 (13.2) | 0.649 |
| Wave 1 | 60.2 (14.3) | 59.6 (19.9) | 0.837 |
| Wave 3 | 56.1 (12.6) | 57.8 (13.2) | 0.383 |
| Wave 5 | 55.9 (11.7) | 58.4 (12.6) | 0.200 |
| **LDL cholesterol (mg/dL), mean (SD)** | | | |
| Baseline | 133.3 (41.0) | 133.1 (28.5) | 0.971 |
| Wave 1 | 127.9 (32.1) | 137.0 (32.1) | 0.074 |
| Wave 3 | 124.6 (31.6) | 133.4 (41.3) | 0.156 |
| Wave 5 | 110.0 (35.5) | 118.8 (42.7) | 0.179 |
| **Systolic blood pressure (mmHg), mean (SD)** | | | |
| Baseline | 131.7 (19.3) | 134.5 (21.2) | 0.154 |
| Wave 1 | 129.7 (18.1) | 130.6 (17.4) | 0.567 |
| Wave 3 | 131.2 (18.7) | 133.1 (18.8) | 0.283 |
| **Diastolic blood pressure (mmHg), mean (SD)** | | | |
| Baseline | 82.3 (10.5) | 83.7 (10.5) | 0.177 |
| Wave 1 | 78.4 (11.3) | 78.9 (10.4) | 0.629 |
| Wave 3 | 80.0 (9.7) | 81.6 (9.9) | 0.106 |
| **Current tobacco use, n (%)** | | | |
| Baseline | 113 (24.5) | 37 (23.4) | 0.792 |
| Wave 1 | 100 (21.6) | 36 (22.8) | 0.765 |
| Wave 3 | 60 (13.0) | 20 (12.7) | 0.915 |
| Wave 5 | 56 (12.1) | 19 (12.0) | 0.975 |
| **Current alcohol use, n (%)** | | | |
| Baseline | 348 (77.0) | 112 (72.3) | 0.235 |
| Wave 1 | 376 (83.2) | 120 (77.4) | 0.109 |
| Wave 3 | 381 (84.3) | 128 (82.6) | 0.617 |
| **Nightly sleep duration (h), mean (SD)** | | | |
| Baseline | 7.5 (1.2) | 7.3 (1.1) | 0.103 |
| Wave 1 | 7.3 (1.0) | 7.4 (1.2) | 0.460 |
| Wave 3 | 6.9 (1.3) | 6.7 (1.2) | 0.057 |
| Wave 5 | 6.8 (1.3) | 6.7 (1.3) | 0.390 |

*Continuous outcomes as mean (SD); binary outcomes as n (%). p-values from Welch's t-tests (continuous outcomes) and Pearson's chi-squared tests without continuity correction (binary outcomes). Wave 5 was not available for blood pressure or alcohol use. Wave 2 was excluded from the primary analysis.*

**Supplementary Table S7.** Pre-test of the parallel-trends assumption for the IPCW analysis.

| Outcome | Adjusted model (*p*-value) | Unadjusted model (*p*-value) |
|---|---|---|
| Body mass index (kg/m²) | 0.971 | 0.874 |
| Overweight/obesity | 0.334 | 0.252 |
| Current tobacco use | 0.033† | 0.029 |
| Current alcohol use | 0.373 | 0.874 |
| Total cholesterol (mg/dL) | 0.051 | 0.177 |
| HDL cholesterol (mg/dL) | 0.405 | 0.465 |
| LDL cholesterol (mg/dL) | 0.024† | 0.116 |
| Systolic blood pressure (mmHg) | 0.009† | <0.001† |
| Diastolic blood pressure (mmHg) | 0.880 | 0.263 |
| Nightly sleep duration (hours) | 0.217 | 0.630 |

*† p < 0.05 indicates a violation of the parallel-trends assumption, in which case the corresponding ATT is not interpretable as a causal effect. Pre-exposure period: baseline and wave 1.*

**Supplementary Table S8.** Wave-specific ATTs of the IPCW analysis (adjusted and unadjusted models).

| Outcome / wave | Adjusted model | | Unadjusted model | |
|---|---|---|---|---|
| | ATT | 95% CI | ATT | 95% CI |
| **Body mass index** | | | | |
| Wave 1 | 0.007 | (−0.369, 0.383) | −0.030 | (−0.387, 0.327) |
| Wave 3 | 0.101 | (−0.548, 0.750) | 0.071 | (−0.642, 0.784) |
| Wave 5 | −0.270 | (−1.219, 0.679) | −0.114 | (−0.974, 0.746) |
| **Overweight/obesity** | | | | |
| Wave 1 | −0.020 | (−0.065, 0.025) | −0.024 | (−0.065, 0.017) |
| Wave 3 | 0.040 | (−0.042, 0.122) | 0.016 | (−0.060, 0.092) |
| Wave 5 | −0.028 | (−0.157, 0.101) | −0.007 | (−0.107, 0.093) |
| **Current tobacco use** | | | | |
| Wave 1 | **0.032** | **(0.003, 0.061)** | **0.029** | **(0.002, 0.056)** |
| Wave 3 | 0.011 | (−0.032, 0.054) | 0.026 | (−0.019, 0.071) |
| Wave 5 | −0.016 | (−0.061, 0.029) | 0.024 | (−0.031, 0.079) |
| **Current alcohol use** | | | | |
| Wave 1 | 0.022 | (−0.027, 0.071) | 0.004 | (−0.037, 0.045) |
| Wave 3 | −0.054 | (−0.130, 0.021) | −0.050 | (−0.126, 0.025) |
| **Total cholesterol** | | | | |
| Wave 1 | 4.784 | (−0.073, 9.641) | 3.260 | (−1.164, 7.684) |
| Wave 3 | −0.005 | (−7.639, 7.629) | −2.693 | (−9.606, 4.220) |
| Wave 5 | −17.336 | (−41.558, 6.886) | 5.202 | (−10.525, 20.929) |
| **HDL cholesterol** | | | | |
| Wave 1 | −0.602 | (−2.031, 0.827) | −0.523 | (−1.858, 0.812) |
| Wave 3 | 1.353 | (−0.795, 3.501) | 0.835 | (−1.213, 2.883) |
| Wave 5 | −1.654 | (−9.986, 6.678) | 1.677 | (−2.294, 5.648) |
| **LDL cholesterol** | | | | |
| Wave 1 | **5.102** | **(0.680, 9.525)** | 3.471 | (−1.208, 8.150) |
| Wave 3 | 0.979 | (−5.415, 7.373) | −1.381 | (−7.136, 4.374) |
| Wave 5 | −10.940 | (−31.381, 9.501) | 5.080 | (−9.018, 19.178) |
| **Systolic blood pressure** | | | | |
| Wave 1 | **−3.576** | **(−6.273, −0.879)** | **−5.832** | **(−8.622, −3.043)** |
| Wave 3 | 2.179 | (−1.768, 6.125) | 1.857 | (−2.057, 5.771) |
| **Diastolic blood pressure** | | | | |
| Wave 1 | 0.126 | (−1.422, 1.674) | −0.920 | (−2.517, 0.677) |
| Wave 3 | 1.072 | (−1.236, 3.379) | 0.955 | (−1.189, 3.099) |
| **Nightly sleep duration** | | | | |
| Wave 1 | 0.127 | (−0.065, 0.319) | 0.048 | (−0.138, 0.234) |
| Wave 3 | **−0.398** | **(−0.714, −0.082)** | **−0.483** | **(−0.781, −0.185)** |
| Wave 5 | −0.294 | (−0.717, 0.129) | **−0.381** | **(−0.691, −0.071)** |

*ATT, average treatment effect on the treated; CI, confidence interval; Wave 5 was not available for blood pressure or alcohol use. Bold denotes statistically significant results.*

**Supplementary Table S9.** Pre-test of the parallel-trends assumption for the complete-case analysis (adjusted and unadjusted models).

| Outcome | Adjusted model (p-value) | Unadjusted model (p-value) |
|---|---|---|
| Body mass index | 0.198 | 0.114 |
| Overweight/obesity | 0.321 | 0.293 |
| Current tobacco use | 0.231 | 0.327 |
| Current alcohol use | 0.864 | 0.805 |
| Total cholesterol | 0.089 | 0.147 |
| HDL cholesterol | 0.947 | 0.873 |
| LDL cholesterol | 0.061 | 0.100 |
| Systolic blood pressure | 0.546 | 0.332 |
| Diastolic blood pressure | 0.477 | 0.447 |
| Nightly sleep duration | 0.030† | 0.032† |

*† $p < 0.05$ indicates a violation of the parallel-trends assumption, in which case the corresponding ATT is not interpretable as a causal effect. Pre-exposure period: baseline and wave 1.*

**Supplementary Table S10.** Wave-specific ATTs for the complete-case analysis (adjusted and unadjusted models).

| Outcome / wave | Adjusted model | | Unadjusted model | |
|---|---|---|---|---|
| | ATT | 95% CI | ATT | 95% CI |
| **Body mass index** | | | | |
| Wave 1 | 0.224 | (−0.169, 0.618) | 0.266 | (−0.133, 0.665) |
| Wave 3 | −0.044 | (−0.622, 0.533) | −0.084 | (−0.643, 0.475) |
| Wave 5 | −0.506 | (−1.284, 0.272) | −0.606 | (−1.397, 0.185) |
| **Overweight/obesity** | | | | |
| Wave 1 | −0.029 | (−0.099, 0.041) | −0.030 | (−0.095, 0.035) |
| Wave 3 | 0.012 | (−0.077, 0.101) | 0.004 | (−0.081, 0.090) |
| Wave 5 | −0.033 | (−0.135, 0.068) | −0.022 | (−0.129, 0.084) |
| **Current tobacco use** | | | | |
| Wave 1 | 0.028 | (−0.025, 0.082) | 0.022 | (−0.030, 0.074) |
| Wave 3 | −0.031 | (−0.093, 0.032) | −0.015 | (−0.078, 0.049) |
| Wave 5 | −0.039 | (−0.106, 0.029) | −0.012 | (−0.079, 0.055) |
| **Current alcohol use** | | | | |
| Wave 1 | 0.007 | (−0.082, 0.096) | −0.010 | (−0.106, 0.086) |
| Wave 3 | 0.026 | (−0.054, 0.105) | 0.041 | (−0.039, 0.120) |
| **Total cholesterol** | | | | |
| Wave 1 | 11.317 | (−5.098, 27.732) | 8.946 | (−5.608, 23.500) |
| Wave 3 | 1.912 | (−14.498, 18.323) | −1.618 | (−19.122, 15.887) |
| Wave 5 | 5.375 | (−11.806, 22.557) | 0.874 | (−17.222, 18.970) |
| **HDL cholesterol** | | | | |
| Wave 1 | 0.172 | (−5.236, 5.579) | 0.419 | (−4.937, 5.774) |
| Wave 3 | 2.704 | (−3.123, 8.532) | 2.364 | (−3.185, 7.913) |
| Wave 5 | 3.545 | (−1.835, 8.925) | 3.068 | (−2.347, 8.483) |
| **LDL cholesterol** | | | | |
| Wave 1 | 11.626 | (−2.596, 25.847) | 9.310 | (−4.066, 22.687) |
| Wave 3 | 2.598 | (−12.088, 17.284) | −0.347 | (−14.993, 14.300) |
| Wave 5 | 3.342 | (−12.907, 19.590) | −0.402 | (−17.104, 16.299) |
| **Systolic blood pressure** | | | | |
| Wave 1 | −1.160 | (−5.581, 3.262) | −1.846 | (−5.981, 2.289) |
| Wave 3 | 0.906 | (−2.575, 4.387) | 0.955 | (−2.442, 4.351) |
| **Diastolic blood pressure** | | | | |
| Wave 1 | −0.808 | (−3.238, 1.622) | −0.855 | (−3.431, 1.721) |
| Wave 3 | 1.069 | (−1.171, 3.309) | 1.024 | (−0.981, 3.028) |
| **Nightly sleep duration** | | | | |
| Wave 1 | 0.262 | (−0.020, 0.543) | 0.250 | (−0.036, 0.537) |
| Wave 3 | −0.307 | (−0.637, 0.022) | −0.299 | (−0.610, 0.012) |
| Wave 5 | −0.196 | (−0.603, 0.212) | −0.189 | (−0.542, 0.165) |

*ATT, average treatment effect on the treated; CI, confidence interval. Wave 5 was not available for blood pressure or alcohol use. Wave 1 is a pre-exposure contrast. Wave 2 was excluded from the primary analysis.*

**Supplementary Table S11.** Final aggregate ATT of the complete-case analysis (adjusted model).

| Outcome | ATT | 95% CI |
|---|---|---|
| Body mass index(kg/m²) | −0.275 | (−0.774, 0.223) |
| Overweight/obesity (proportion) | −0.011 | (−0.086, 0.064) |
| Total cholesterol (mg/dL) | 3.644 | (−9.453, 16.741) |
| HDL cholesterol (mg/dL) | 3.125 | (−2.709, 8.959) |
| LDL cholesterol (mg/dL) | 2.970 | (−10.314, 16.255) |
| Systolic blood pressure (mmHg) | 0.906 | (−2.575, 4.387) |
| Diastolic blood pressure (mmHg) | 1.069 | (−1.171, 3.309) |
| Current tobacco use (proportion) | −0.035 | (−0.090, 0.020) |
| Current alcohol use (proportion) | 0.026 | (−0.054, 0.105) |
| Nightly sleep duration (hours) | −0.252 | (−0.509, 0.005) |

*ATT, average treatment effect on the treated;*

**Supplementary Table S12.** Final aggregate ATT of the complete-case analysis (unadjusted model).

| Outcome | ATT | 95% CI |
|---|---|---|
| Body mass index (kg/m²) | −0.345 | (−0.873, 0.183) |
| Overweight/obesity (proportion) | −0.009 | (−0.079, 0.061) |
| Total cholesterol (mg/dL) | −0.372 | (−13.891, 13.148) |
| HDL cholesterol (mg/dL) | 2.716 | (−2.662, 8.094) |
| LDL cholesterol (mg/dL) | −0.375 | (−12.476, 11.727) |
| Systolic blood pressure (mmHg) | 0.955 | (−2.442, 4.351) |
| Diastolic blood pressure (mmHg) | 1.024 | (−0.981, 3.028) |
| Current tobacco use (proportion) | −0.014 | (−0.066, 0.039) |
| Current alcohol use (proportion) | 0.041 | (−0.039, 0.120) |
| Nightly sleep duration (hours) | −0.244 | (−0.513, 0.025) |

*ATT, average treatment effect on the treated*

### Section C. Cluster-bootstrap assessment of the stability of complete-case estimates

**Supplementary Table S13.** Cluster-bootstrap estimates and sign consistency across 2,000 and 5,000 replicates (complete-case analysis).

| Outcome | 2,000 replicates | | | 5,000 replicates | | |
|---|---|---|---|---|---|---|
| | Bootstrap mean | 95% CI | Sign consistency | Bootstrap mean | 95% CI | Sign consistency |
| BMI (kg/m²) | −0.234 | (−0.948, 0.477) | 0.74 | −0.239 | (−0.963, 0.481) | 0.74 |
| Overweight/obesity | −0.006 | (−0.106, 0.097) | 0.54 | −0.004 | (−0.102, 0.094) | 0.53 |
| Total cholesterol | 4.074 | (−16.132, 25.274) | 0.65 | 4.066 | (−16.118, 23.854) | 0.66 |
| HDL cholesterol | 3.100 | (−3.926, 9.303) | 0.79 | 3.138 | (−3.919, 9.327) | 0.80 |
| LDL cholesterol | 3.709 | (−14.698, 22.210) | 0.66 | 3.399 | (−15.820, 22.120) | 0.65 |
| Systolic blood pressure | 0.911 | (−4.121, 5.745) | 0.66 | 0.912 | (−4.065, 5.614) | 0.65 |
| Diastolic blood pressure | 1.085 | (−1.813, 4.198) | 0.76 | 1.051 | (−1.934, 4.105) | 0.76 |
| Tobacco use | −0.040 | (−0.117, 0.030) | 0.86 | −0.040 | (−0.113, 0.031) | 0.86 |
| Alcohol use | 0.031 | (−0.083, 0.145) | 0.71 | 0.030 | (−0.083, 0.146) | 0.70 |
| Nightly sleep duration | −0.270 | (−0.639, 0.097) | 0.93 | −0.266 | (−0.632, 0.107) | 0.92 |

*Sign consistency is the proportion of bootstrap replicates yielding an estimate of the same sign as the point estimate; by construction it approaches 0.5 when the direction of the estimate is unstable and does not represent statistical power.*

### Section D. Subgroup analyses by educational attainment and age

**Supplementary Table S14.** Pre-test of the parallel-trends assumption for the complete-case analysis within subgroups (unadjusted models).

| Outcome | **Lower education** *(p-value)* | **Higher education** *(p-value)* | **≥65 years** *(p-value)* | **<65 years** *(p-value)* |
|---|---|---|---|---|
| Body mass index (kg/m²) | 0.222 | 0.271 | 0.196 | 0.326 |
| Overweight/obesity | 0.226 | 0.282 | 0.219 | 0.847 |
| Total cholesterol (mg/dL) | 0.017† | 0.789 | 0.070 | 0.801 |
| HDL cholesterol (mg/dL) | 0.378 | 0.442 | 0.940 | 0.842 |
| LDL cholesterol (mg/dL) | 0.037† | 0.899 | 0.012† | 0.595 |
| Systolic blood pressure (mmHg) | 0.460 | 0.136 | 0.360 | 0.599 |
| Diastolic blood pressure (mmHg) | 0.284 | 0.351 | 0.784 | 0.825 |
| Current tobacco use | 0.498 | 0.506 | 0.885 | 0.099 |
| Current alcohol use | 0.967 | 0.554 | 0.887 | 0.937 |
| Nightly sleep duration (hours) | 0.062 | 0.894 | 0.184 | 0.160 |

*Education: lower education (≤9 years, ISCED 0–2), higher education (>12 years, ISCED 5–8). † $p < 0.05$ indicates a violation of the parallel-trends assumption, in which case the corresponding ATT is not interpretable as a causal effect.*

**Supplementary Table S15.** Pre-test of the parallel-trends assumption for the complete-case analysis within subgroups (adjusted models).

| Outcome | **Lower education** *(p-value)* | **Higher education** *(p-value)* | **≥65 years** *(p-value)* | **<65 years** *(p-value)* |
|---|---|---|---|---|
| Body mass index (kg/m²) | 0.496 | 0.189 | 0.367 | 0.318 |
| Overweight/obesity | 0.318 | 0.224 | 0.210 | 0.909 |
| Total cholesterol (mg/dL) | 0.118 | 0.966 | 0.038† | 0.455 |
| HDL cholesterol (mg/dL) | 0.466 | 0.783 | 0.830 | 0.663 |
| LDL cholesterol (mg/dL) | 0.101 | 0.761 | 0.007† | 0.918 |
| Systolic blood pressure (mmHg) | 0.060 | 0.102 | 0.488 | 0.468 |
| Diastolic blood pressure (mmHg) | 0.650 | 0.538 | 0.681 | 0.854 |
| Current tobacco use | 0.120 | 0.667 | 0.978 | 0.086 |
| Current alcohol use | 0.172 | 0.520 | 0.740 | 0.723 |
| Nightly sleep duration (hours) | 0.231 | 0.787 | 0.255 | 0.130 |

*Education: lower education (≤9 years, ISCED 0–2), higher education (>12 years, ISCED 5–8). † $p < 0.05$ indicates a violation of the parallel-trends assumption, in which case the corresponding ATT is not interpretable as a causal effect.*

**Supplementary Table S16**. Final aggregate ATT for the complete-case analysis within subgroups (unadjusted models).

| Outcome | **Lower education** ATT (95% CI) | **Higher education** ATT (95% CI) | **Older (≥65 years)** ATT (95% CI) | **Younger (<65 years)** ATT (95% CI) |
|---|---|---|---|---|
| Body mass index (kg/m²) | −0.284 (−1.117, 0.548) | 0.726 (−0.343, 1.796) | −0.308 (−0.974, 0.359) | −0.237 (−1.129, 0.654) |
| Overweight/obesity | 0.011 (−0.094, 0.116) | 0.073 (−0.068, 0.213) | 0.018 (−0.072, 0.109) | −0.038 (−0.166, 0.091) |
| Total cholesterol (mg/dL) | 3.081 (−15.377, 21.538) | −3.655 (−22.173, 14.863) | 1.017 (−14.299, 16.333) | 0.088 (−21.740, 21.915) |
| HDL cholesterol (mg/dL) | 4.155 (−0.682, 8.992) | 2.461 (−1.627, 6.549) | 2.918 (−1.113, 6.949) | 2.113 (−9.052, 13.277) |
| LDL cholesterol (mg/dL) | 6.969 (−10.165, 24.102) | −5.396 (−20.786, 9.993) | −1.689 (−16.299, 12.922) | 4.009 (−13.147, 21.164) |
| Systolic blood pressure (mmHg) | 1.251 (−3.728, 6.230) | −3.681 (−10.825, 3.464) | −0.950 (−5.443, 3.543) | 3.580 (−3.199, 10.359) |
| Diastolic blood pressure (mmHg) | 1.296 (−2.482, 5.074) | 2.648 (−2.132, 7.428) | 0.304 (−2.557, 3.166) | 2.430 (−3.042, 7.903) |
| Current tobacco use | −0.039 (−0.109, 0.031) | 0.079 (−0.045, 0.202) | 0.000 (−0.058, 0.058) | −0.036 (−0.157, 0.085) |
| Current alcohol use | 0.008 (−0.110, 0.127) | 0.041 (−0.102, 0.184) | 0.012 (−0.099, 0.123) | 0.053 (−0.063, 0.168) |
| Nightly sleep duration (hours) | −0.391 (−0.797, 0.015) | 0.099 (−0.290, 0.489) | −0.233 (−0.549, 0.083) | −0.196 (−0.665, 0.273) |

*Education: lower education (≤9 years, ISCED 0–2), higher education (>12 years, ISCED 5–8). ATT, average treatment effect on the treated; CI, confidence interval;*

**Supplementary Table S17.** Final aggregate ATT for the complete-case analysis within subgroups (adjusted models).

| Outcome | **Lower education** ATT (95% CI) | **Higher education** ATT (95% CI) | **Older (≥65 years)** ATT (95% CI) | **Younger (<65 years)** ATT (95% CI) |
|---|---|---|---|---|
| Body mass index (kg/m²) | −0.313 (−1.143, 0.516) | 0.540 (−0.436, 1.517) | −0.259 (−0.929, 0.410) | −0.186 (−1.094, 0.721) |
| Overweight/obesity | 0.017 (−0.073, 0.107) | 0.056 (−0.081, 0.193) | 0.015 (−0.065, 0.094) | −0.043 (−0.169, 0.083) |
| Total cholesterol (mg/dL) | 1.077 (−19.178, 21.332) | 5.750 (−10.836, 22.335) | 2.339 (−15.826, 20.503) | 3.689 (−18.569, 25.947) |
| HDL cholesterol (mg/dL) | 4.834 (−0.065, 9.733) | 0.803 (−3.742, 5.347) | 2.216 (−1.804, 6.235) | 2.621 (−8.235, 13.477) |
| LDL cholesterol (mg/dL) | 5.060 (−11.843, 21.963) | 1.994 (−12.282, 16.270) | −0.735 (−20.298, 18.828) | 5.809 (−10.609, 22.226) |
| Systolic blood pressure (mmHg) | 1.628 (−3.866, 7.122) | 2.567 (−1.613, 6.747) | −0.552 (−4.967, 3.863) | 2.970 (−4.376, 10.317) |
| Diastolic blood pressure (mmHg) | 1.318 (−2.219, 4.855) | 0.232(−5.936, 6.400) | 0.783 (−2.090, 3.655) | 2.177 (−4.272, 8.625) |
| Current tobacco use | −0.044 (−0.112, 0.024) | 0.115 (−0.003, 0.232) | −0.008 (−0.070, 0.054) | −0.056 (−0.176, 0.064) |
| Current alcohol use | −0.014 (−0.133, 0.106) | 0.037 (−0.091, 0.166) | −0.004 (−0.112, 0.103) | 0.036 (−0.083, 0.154) |
| Nightly sleep duration (hours) | −0.351 (−0.766, 0.063) | 0.134 (−0.303, 0.572) | −0.188 (−0.529, 0.154) | −0.269 (−0.754, 0.216) |

*Education: lower education (≤9 years, ISCED 0–2), higher education (>12 years, ISCED 5–8). ATT, average treatment effect on the treated; CI, confidence interval;*

**Supplementary Table S18.** Pre-test of the parallel-trends assumption for the IPCW analysis within subgroups (unadjusted models).

| **Outcome** | **Lower education** *(p-value)* | **Higher education** *(p-value)* | **Older (≥65 years)** *(p-value)* | **Younger (<65 years)** *(p-value)* |
|---|---|---|---|---|
| Body mass index (kg/m²) | 0.691 | 0.230 | 0.747 | 0.550 |
| Overweight/obesity | 0.234 | 0.220 | 0.179 | 0.682 |
| Total cholesterol (mg/dL) | 0.174 | 0.891 | 0.213 | 0.087 |
| HDL cholesterol (mg/dL) | 0.516 | 0.372 | 0.177 | 0.568 |
| LDL cholesterol (mg/dL) | 0.160 | 0.771 | 0.071 | 0.215 |
| Systolic blood pressure (mmHg) | 0.013† | 0.139 | 0.001† | 0.101 |
| Diastolic blood pressure (mmHg) | 0.946 | 0.279 | 0.923 | 0.123 |
| Current tobacco use | 0.090 | 0.591 | 0.281 | 0.100 |
| Current alcohol use | 0.217 | 0.515 | 0.365 | 0.446 |
| Nightly sleep duration (hours) | 0.373 | 0.801 | 0.452 | 0.937 |

*Education: lower education (≤9 years, ISCED 0–2), higher education (>12 years, ISCED 5–8). † $p < 0.05$ indicates a violation of the parallel-trends assumption, in which case the corresponding ATT is not interpretable as a causal effect.*

**Supplementary Table S19.** Pre-test of the parallel-trends assumption for the IPCW analysis within subgroups (adjusted models).

| **Outcome** | **Lower education** *(p-value)* | **Higher education** *(p-value)* | **Older (≥65 years)** *(p-value)* | **Younger (<65 years)** *(p-value)* |
|---|---|---|---|---|
| Body mass index (kg/m²) | 0.551 | 0.188 | 0.693 | 0.667 |
| Overweight/obesity | 0.280 | 0.224 | 0.150 | 0.958 |
| Total cholesterol (mg/dL) | 0.109 | 0.983 | 0.148 | 0.108 |
| HDL cholesterol (mg/dL) | 0.459 | 0.771 | 0.217 | 0.389 |
| LDL cholesterol (mg/dL) | 0.093 | 0.718 | 0.049† | 0.232 |
| Systolic blood pressure (mmHg) | 0.051 | 0.346 | 0.015† | 0.285 |
| Diastolic blood pressure (mmHg) | 0.693 | 0.554 | 0.541 | 0.642 |
| Current tobacco use | 0.106 | 0.750 | 0.155 | 0.041† |
| Current alcohol use | 0.158 | 0.471 | 0.137 | 0.647 |
| Nightly sleep duration (hours) | 0.213 | 0.736 | 0.170 | 0.999 |

*Education: lower education (≤9 years, ISCED 0–2), higher education (>12 years, ISCED 5–8). † $p < 0.05$ indicates a violation of the parallel-trends assumption, in which case the corresponding ATT is not interpretable as a causal effect.*

**Supplementary Table S20.** Final aggregate ATT for the IPCW analysis within subgroups (unadjusted models)

| Outcome | Lower education<br>ATT (95% CI) | Higher education<br>ATT (95% CI) | Older (≥65 years)<br>ATT (95% CI) | Younger (<65 years)<br>ATT (95% CI) |
|---|---|---|---|---|
| Body mass index (kg/m²) | 0.176 (−0.763, 1.116) | 0.701 (−0.391, 1.793) | −0.171 (−1.032, 0.690) | 0.932 (−0.045, 1.908) |
| Overweight/obesity | 0.023 (−0.083, 0.129) | 0.072 (−0.078, 0.222) | 0.007 (−0.089, 0.102) | 0.042 (−0.081, 0.165) |
| Total cholesterol (mg/dL) | 3.307 (−9.620, 16.234) | −4.553 (−22.790, 13.684) | 0.395 (−12.204, 12.995) | 4.980 (−8.954, 18.915) |
| HDL cholesterol (mg/dL) | 1.514 (−2.455, 5.484) | 2.105 (−1.910, 6.119) | 0.503 (−3.091, 4.097) | 1.964 (−1.969, 5.896) |
| LDL cholesterol (mg/dL) | 4.828 (−7.182, 16.837) | −6.284 (−20.083, 7.514) | 2.140 (−10.010, 14.290) | 4.370 (−9.003, 17.742) |
| Systolic blood pressure (mmHg) | 2.024 (−3.176, 7.225) | −3.608 (−10.960, 3.744) | 1.779 (−2.777, 6.335) | 3.498 (−2.431, 9.427) |
| Diastolic blood pressure (mmHg) | 0.198 (−2.534, 2.929) | 3.068 (−1.736, 7.871) | 1.005 (−1.432, 3.441) | 1.872 (−2.721, 6.465) |
| Current tobacco use | 0.017 (−0.034, 0.067) | 0.095 (−0.035, 0.224) | 0.012 (−0.031, 0.055) | 0.001 (−0.105, 0.107) |
| Current alcohol use | −0.078 (−0.177, 0.020) | 0.062 (−0.076, 0.200) | −0.072 (−0.165, 0.020) | 0.032 (−0.064, 0.128) |
| Nightly sleep duration (hours) | **−0.475 (−0.804, −0.146)** | 0.121 (−0.308, 0.550) | **−0.491 (−0.819, −0.164)** | −0.131 (−0.501, 0.239) |

*Education: lower education (≤9 years, ISCED 0–2), higher education (>12 years, ISCED 5–8); ATT, average treatment effect on the treated; CI, confidence interval; Bold denotes statistically significant results.*

### Section E. Sensitivity analysis re-including follow-up Wave 2

In the primary analysis, wave 2 was excluded. As a sensitivity analysis, this wave was re-included and treated as the first post-exposure wave, so that the post-exposure period comprises follow-up waves 2, 3 and 5, when available. Estimates are reported for the complete-case and the IPCW analysis, considering both the unadjusted and the adjusted model, alongside with the corresponding parallel-trends pre-tests. Nightly sleep duration was not assessed at wave 2 and could not therefore be included in this sensitivity analysis.

**Supplementary Table S21.** Pre-test of the parallel-trends assumption for the complete-case analysis for the sensitivity analysis including wave 2 (adjusted and unadjusted models).

| Outcome | Adjusted model (p-value) | Unadjusted model (p-value) |
|---|---|---|
| Body mass index (kg/m²) | 0.595 | 0.410 |
| Overweight/obesity | 0.179 | 0.222 |
| Current tobacco use | 0.153 | 0.245 |
| Current alcohol use | 0.756 | 0.956 |
| Total cholesterol (mg/dL) | 0.174 | 0.197 |
| HDL cholesterol (mg/dL) | 0.503 | 0.518 |
| LDL cholesterol (mg/dL) | 0.145 | 0.164 |
| Systolic blood pressure (mmHg) | 0.899 | 0.526 |
| Diastolic blood pressure (mmHg) | 0.721 | 0.486 |
| Nightly sleep duration (hours) | — | — |

*Pre-exposure period: baseline and wave 1.*

**Supplementary Table S22**. Final aggregate ATT for the complete-case analysis for the sensitivity analysis including wave 2 (adjusted and unadjusted models).

| Outcome | Adjusted ATT (95% CI) | Unadjusted ATT (95% CI) |
|---|---|---|
| Body mass index (kg/m²) | −0.245 (−0.734, 0.244) | −0.262 (−0.740, 0.215) |
| Overweight/obesity | −0.015 (−0.088, 0.058) | −0.013 (−0.086, 0.059) |
| Current tobacco use | −0.011 (−0.066, 0.045) | 0.004 (−0.052, 0.061) |
| Current alcohol use | −0.001 (−0.075, 0.074) | 0.006 (−0.067, 0.080) |
| Total cholesterol (mg/dL) | 3.149 (−8.700, 14.997) | −0.267 (−13.057, 12.522) |
| HDL cholesterol (mg/dL) | 1.656 (−3.594, 6.906) | 1.395 (−3.795, 6.585) |
| LDL cholesterol (mg/dL) | 2.070 (−8.659, 12.800) | −1.566 (−12.683, 9.552) |
| Systolic blood pressure (mmHg) | 2.504 (−0.870, 5.877) | 2.317 (−1.000, 5.635) |
| Diastolic blood pressure (mmHg) | 1.086 (−1.013, 3.184) | 0.928 (−1.091, 2.947) |
| Nightly sleep duration (hours) | — | — |

*ATT, average treatment effect on the treated; CI, confidence interval;*

**Supplementary Table S23.** Pre-test of the parallel-trends assumption for the IPCW analysis for the sensitivity analysis including wave 2 (adjusted and unadjusted models).

| Outcome | Adjusted model (p-value) | Unadjusted model (p-value) |
|---|---|---|
| Body mass index (kg/m²) | 0.967 | 0.910 |
| Overweight/obesity | 0.336 | 0.276 |
| Current tobacco use | 0.335 | 0.065 |
| Current alcohol use | 0.368 | 0.867 |
| Total cholesterol (mg/dL) | 0.055 | 0.183 |
| HDL cholesterol (mg/dL) | 0.400 | 0.462 |
| LDL cholesterol (mg/dL) | 0.205 | 0.122 |
| Systolic blood pressure (mmHg) | 0.319 | 0.061 |
| Diastolic blood pressure (mmHg) | 0.891 | 0.255 |
| Nightly sleep duration (hours) | — | — |

*Pre-exposure period: baseline and wave 1.*

**Supplementary Table S24.** Final aggregate ATT for the IPCW analysis for the sensitivity analysis including wave 2 (adjusted and unadjusted models).

| Outcome | Adjusted ATT (95% CI) | Unadjusted ATT (95% CI) |
|---|---|---|
| Body mass index (kg/m²) | −0.164 (−0.835, 0.507) | −0.056 (−0.708, 0.596) |
| Overweight/obesity | −0.030 (−0.107, 0.048) | −0.016 (−0.085, 0.053) |
| Current tobacco use | 0.013 (−0.028, 0.053) | 0.041 (−0.002, 0.085) |
| Current alcohol use | −0.058 (−0.129, 0.013) | −0.043 (−0.107, 0.022) |
| Total cholesterol (mg/dL) | −10.742 (−21.950, 0.466) | −0.809 (−8.654, 7.035) |
| HDL cholesterol (mg/dL) | −1.190 (−4.851, 2.471) | 0.590 (−1.563, 2.742) |
| LDL cholesterol (mg/dL) | −8.275 (−17.816, 1.266) | −0.109 (−7.280, 7.062) |
| Systolic blood pressure (mmHg) | 2.405 (−0.925, 5.734) | 1.247 (−2.258, 4.752) |
| Diastolic blood pressure (mmHg) | 0.780 (−1.156, 2.717) | 0.378 (−1.456, 2.211) |
| Nightly sleep duration (hours) | — | — |

*ATT, average treatment effect on the treated; CI, confidence interval;*

### Section F. Sensitivity analyses using alternative cut-off points

**Supplementary Table S25.** Alternative cut-off points for exposure definition.

| Cut-off point | Number of census block groups considered gentrifying | Value of the index at the cut-off point |
|---|---|---|
| 1 SD above the mean | 144 | 1.000 |
| 1.5 SD above the mean | 87 | 1.500 |
| Upper quartile | 361 | 0.401 |

*SD, standard deviation;*

**Supplementary Table S26.** Pre-test of the parallel-trends assumption for the complete-case analysis (adjusted and unadjusted models) under the alternative exposure definitions.

| Outcome | ≥1.5 SD above the mean | | Upper quartile of the index | |
|---|---|---|---|---|
| | Adjusted (p) | Unadjusted (p) | Adjusted (p) | Unadjusted (p) |
| Body mass index (kg/m²) | 0.435 | 0.455 | 0.618 | 0.812 |
| Overweight/obesity | 0.834 | 0.832 | 0.567 | 0.667 |
| Current tobacco use | 0.171 | 0.199 | 0.293 | 0.261 |
| Current alcohol use | 0.077 | 0.155 | 0.528 | 0.307 |
| Total cholesterol (mg/dL) | 0.365 | 0.348 | 0.978 | 0.699 |
| HDL cholesterol (mg/dL) | 0.964 | 0.864 | 0.827 | 0.879 |
| LDL cholesterol (mg/dL) | 0.516 | 0.545 | 0.915 | 0.762 |
| Systolic blood pressure (mmHg) | 0.289 | 0.115 | 0.537 | 0.242 |
| Diastolic blood pressure (mmHg) | 0.074 | 0.135 | 0.228 | 0.160 |
| Nightly sleep duration (hours) | 0.104 | 0.100 | 0.320 | 0.420 |

*Pre-exposure period: baseline and wave 1.*

**Supplementary Table S27.** Final aggregate ATT for the complete-case analysis (adjusted and unadjusted models) under the alternative exposure definitions.

| Outcome | ≥1.5 SD above the mean | | Upper quartile of the index | |
|---|---|---|---|---|
| | Adjusted ATT (95% CI) | Unadjusted ATT (95% CI) | Adjusted ATT (95% CI) | Unadjusted ATT (95% CI) |
| Body mass index (kg/m²) | −0.175 (−0.788, 0.439) | −0.385 (−1.043, 0.273) | −0.401 (−0.890, 0.087) | −0.431 (−0.950, 0.088) |
| Overweight/obesity | −0.021 (−0.107, 0.064) | −0.037 (−0.131, 0.056) | 0.022 (−0.044, 0.089) | 0.006 (−0.061, 0.073) |
| Current tobacco use | −0.033 (−0.106, 0.041) | −0.015 (−0.093, 0.062) | −0.026 (−0.070, 0.018) | −0.010 (−0.059, 0.039) |
| Current alcohol use | −0.027 (−0.131, 0.077) | −0.038 (−0.139, 0.064) | 0.034 (−0.043, 0.112) | 0.041 (−0.034, 0.117) |
| Total cholesterol (mg/dL) | 6.830 (−8.686, 22.346) | 1.228 (−16.785, 19.241) | −4.826 (−17.453, 7.801) | −8.374 (−21.499, 4.752) |
| HDL cholesterol (mg/dL) | 3.452 (−0.251, 7.155) | 3.417 (−0.009, 6.843) | 1.837 (−1.593, 5.267) | 1.091 (−2.662, 4.844) |
| LDL cholesterol (mg/dL) | 8.267 (−5.895, 22.430) | 4.122 (−11.269, 19.513) | −4.117 (−16.272, 8.038) | −6.343 (−18.143, 5.457) |
| Systolic blood pressure (mmHg) | 2.566 (−1.662, 6.795) | 2.446 (−2.051, 6.944) | 2.296 (−1.326, 5.919) | 2.459 (−0.873, 5.790) |
| Diastolic blood pressure (mmHg) | 2.447 (−0.884, 5.779) | 2.540 (−0.698, 5.779) | 1.716 (−0.709, 4.141) | 1.555 (−0.388, 3.498) |
| Nightly sleep duration (hours) | −0.129 (−0.445, 0.187) | −0.131 (−0.464, 0.201) | −0.158 (−0.431, 0.115) | −0.183 (−0.427, 0.061) |

*ATT, average treatment effect on the treated; CI, confidence interval;*

**Supplementary Table S28.** Pre-test of the parallel-trends assumption for the IPCW analysis (adjusted and unadjusted models) under the alternative exposure definitions.

| Outcome | ≥1.5 SD above the mean | | Upper quartile of the index | |
|---|---|---|---|---|
| | **Adjusted (p)** | **Unadjusted (p)** | **Adjusted (p)** | **Unadjusted (p)** |
| Body mass index (kg/m²) | 0.604 | 0.914 | 0.873 | 0.897 |
| Overweight/obesity | 0.878 | 0.636 | 0.598 | 0.566 |
| Current tobacco use | 0.524 | 0.337 | 0.674 | 0.359 |
| Current alcohol use | 0.182 | 0.361 | 0.415 | 0.141 |
| Total cholesterol (mg/dL) | 0.417 | 0.561 | 0.614 | 0.789 |
| HDL cholesterol (mg/dL) | 0.909 | 0.792 | 0.912 | 0.896 |
| LDL cholesterol (mg/dL) | 0.310 | 0.746 | 0.441 | 0.854 |
| Systolic blood pressure (mmHg) | 0.112 | 0.065 | 0.195 | 0.081 |
| Diastolic blood pressure (mmHg) | 0.554 | 0.173 | 0.161 | 0.072 |
| Nightly sleep duration (hours) | 0.110 | 0.435 | 0.279 | 0.687 |

*Pre-exposure period: baseline and wave 1.*

**Supplementary Table S29**. Final aggregate ATT for the IPCW analysis (adjusted and unadjusted models) under the alternative exposure definitions.

| Outcome | ≥1.5 SD above the mean | | Upper quartile of the index | |
|---|---|---|---|---|
| | **Adjusted ATT (95% CI)** | **Unadjusted ATT (95% CI)** | **Adjusted ATT (95% CI)** | **Unadjusted ATT (95% CI)** |
| Body mass index (kg/m²) | −0.765 (−1.800, 0.270) | −0.809 (−1.711, 0.092) | 0.416 (−0.265, 1.098) | −0.064 (−0.674, 0.545) |
| Overweight/obesity | −0.062 (−0.199, 0.075) | −0.066 (−0.165, 0.034) | 0.061 (−0.032, 0.154) | 0.008 (−0.068, 0.084) |
| Current tobacco use | −0.027 (−0.079, 0.026) | −0.013 (−0.072, 0.045) | 0.021 (−0.025, 0.066) | 0.040 (−0.006, 0.086) |
| Current alcohol use | −0.041 (−0.145, 0.064) | −0.057 (−0.160, 0.046) | 0.038 (−0.044, 0.119) | 0.036 (−0.034, 0.106) |
| Total cholesterol (mg/dL) | −9.578 (−23.377, 4.222) | 1.851 (−12.261, 15.964) | −14.709 (−32.617, 3.198) | −9.821 (−20.085, 0.444) |
| HDL cholesterol (mg/dL) | 0.044 (−3.881, 3.969) | 2.515 (−0.491, 5.521) | −0.424 (−3.774, 2.926) | −0.113 (−2.534, 2.307) |
| LDL cholesterol (mg/dL) | −0.874 (−13.432, 11.684) | 5.979 (−5.513, 17.469) | −12.735 (−28.455, 2.985) | −8.583 (−17.379, 0.214) |
| Systolic blood pressure (mmHg) | 4.153 (−0.954, 9.260) | 2.504 (−2.655, 7.662) | 1.672 (−2.311, 5.654) | 2.034 (−1.871, 5.940) |
| Diastolic blood pressure (mmHg) | 2.859 (−0.239, 5.957) | 2.022 (−0.637, 4.681) | 0.720 (−1.707, 3.147) | 1.589 (−0.406, 3.586) |
| Nightly sleep duration (hours) | **−0.349 (−0.658, −0.040)** | **−0.414 (−0.757, −0.071)** | **−0.350 (−0.697, −0.003)** | **−0.443 (−0.692, −0.193)** |

*ATT, average treatment effect on the treated; CI, confidence interval; Bold denotes statistically significant results.*

**Supplementary Table S30.** Pre-test of the parallel-trends assumption for the complete-case analysis within subgroups under the ≥1.5 SD definition (unadjusted models).

| Outcome | **Lower education** *(p-value)* | **Higher education** *(p-value)* | **Older (≥65 years)** *(p-value)* | **Younger (<65 years)** *(p-value)* |
|---|---|---|---|---|
| Body mass index (kg/m²) | 0.534 | 0.597 | 0.948 | 0.927 |
| Overweight/obesity | 0.607 | 0.684 | 0.510 | 0.398 |
| Total cholesterol (mg/dL) | 0.515 | 0.816 | 0.071 | 0.221 |
| HDL cholesterol (mg/dL) | 0.763 | 0.665 | 0.678 | 0.438 |
| LDL cholesterol (mg/dL) | **0.012**† | 0.839 | **0.036**† | 0.355 |
| Systolic blood pressure (mmHg) | 0.050 | 0.425 | **0.009**† | 0.117 |
| Diastolic blood pressure (mmHg) | 0.650 | 0.350 | 0.601 | 0.215 |
| Current tobacco use | 0.220 | 0.964 | 0.076 | 0.250 |
| Current alcohol use | 0.318 | 0.213 | 0.295 | 0.839 |
| Nightly sleep duration (hours) | 0.138 | 0.143 | 0.112 | 0.822 |

*Education: lower education (≤9 years, ISCED 0–2), higher education (>12 years, ISCED 5–8). † p < 0.05 (bold) indicates a violation of the parallel-trends assumption, in which case the corresponding ATT is not interpretable as a causal effect. Pre-exposure period: baseline and wave 1. Within each stratum, the stratifying variable was omitted from the adjustment set because it was constant.*

**Supplementary Table S31.** Pre-test of the parallel-trends assumption for the complete-case analysis within subgroups under the ≥1.5 SD definition (adjusted models).

| **Outcome** | **Lower education** *(p-value)* | **Higher education** *(p-value)* | **Older (≥65 years)** *(p-value)* | **Younger (<65 years)** *(p-value)* |
|---|---|---|---|---|
| Body mass index (kg/m²) | 0.442 | 0.742 | 0.474 | 0.869 |
| Overweight/obesity | 0.577 | 0.531 | 0.683 | 0.413 |
| Total cholesterol (mg/dL) | 0.060 | 0.864 | 0.127 | 0.188 |
| HDL cholesterol (mg/dL) | 0.782 | 0.912 | 0.721 | 0.428 |
| LDL cholesterol (mg/dL) | **0.007**† | 0.407 | **0.015**† | 0.277 |
| Systolic blood pressure (mmHg) | 0.075 | 0.058 | **0.027**† | 0.105 |
| Diastolic blood pressure (mmHg) | 0.871 | 0.176 | 0.737 | 0.455 |
| Current tobacco use | 0.151 | 0.689 | 0.062 | 0.189 |
| Current alcohol use | 0.242 | 0.344 | 0.255 | 0.357 |
| Nightly sleep duration (hours) | 0.099 | 0.812 | 0.375 | 0.682 |

*Education: lower education (≤9 years, ISCED 0–2), higher education (>12 years, ISCED 5–8). † p < 0.05 (bold) indicates a violation of the parallel-trends assumption, in which case the corresponding ATT is not interpretable as a causal effect. Pre-exposure period: baseline and wave 1. Within each stratum, the stratifying variable was omitted from the adjustment set because it was constant.*

**Supplementary Table S32.** Pre-test of the parallel-trends assumption for the IPCW analysis within subgroups under the ≥1.5 SD definition (unadjusted models).

| **Outcome** | **Lower education** *(p-value)* | **Higher education** *(p-value)* | **Older (≥65 years)** *(p-value)* | **Younger (<65 years)** *(p-value)* |
|---|---|---|---|---|
| Body mass index (kg/m²) | 0.535 | 0.467 | 0.845 | 0.962 |
| Overweight/obesity | 0.436 | 0.880 | 0.417 | 0.400 |
| Total cholesterol (mg/dL) | **0.018**† | 0.848 | 0.059 | 0.215 |
| HDL cholesterol (mg/dL) | 0.697 | 0.789 | 0.553 | 0.563 |
| LDL cholesterol (mg/dL) | **0.015**† | 0.913 | **0.028**† | 0.333 |
| Systolic blood pressure (mmHg) | 0.053 | 0.349 | 0.057 | 0.099 |
| Diastolic blood pressure (mmHg) | 0.673 | 0.273 | 0.679 | **0.018**† |
| Current tobacco use | 0.182 | 0.785 | 0.064 | 0.271 |
| Current alcohol use | 0.312 | 0.209 | 0.304 | 0.776 |
| Nightly sleep duration (hours) | 0.128 | 0.168 | **0.013**† | 0.777 |

*Education: lower education (≤9 years, ISCED 0–2), higher education (>12 years, ISCED 5–8). † $p < 0.05$ (bold) indicates a violation of the parallel-trends assumption, in which case the corresponding ATT is not interpretable as a causal effect. Pre-exposure period: baseline and wave 1. Within each stratum, the stratifying variable was omitted from the adjustment set because it was constant.*

**Supplementary Table S33.** Pre-test of the parallel-trends assumption for the IPCW analysis within subgroups under the ≥1.5 SD definition (adjusted models).

| **Outcome** | **Lower education** *(p-value)* | **Higher education** *(p-value)* | **Older (≥65 years)** *(p-value)* | **Younger (<65 years)** *(p-value)* |
|---|---|---|---|---|
| Body mass index (kg/m²) | 0.416 | 0.618 | 0.470 | 0.818 |
| Overweight/obesity | 0.527 | 0.774 | 0.606 | 0.452 |
| Total cholesterol (mg/dL) | **0.009**† | 0.439 | **0.023**† | 0.195 |
| HDL cholesterol (mg/dL) | 0.705 | 0.856 | 0.677 | 0.408 |
| LDL cholesterol (mg/dL) | **0.006**† | 0.592 | **0.013**† | 0.291 |
| Systolic blood pressure (mmHg) | 0.060 | 0.087 | **0.020**† | 0.067 |
| Diastolic blood pressure (mmHg) | 0.805 | 0.231 | 0.792 | 0.322 |
| Current tobacco use | 0.152 | 0.508 | 0.063 | 0.197 |
| Current alcohol use | 0.246 | 0.129 | 0.259 | 0.368 |
| Nightly sleep duration (hours) | 0.081 | 0.963 | **0.005**† | 0.672 |

*Education: lower education (≤9 years, ISCED 0–2), higher education (>12 years, ISCED 5–8). † $p < 0.05$ (bold) indicates a violation of the parallel-trends assumption, in which case the corresponding ATT is not interpretable as a causal effect. Pre-exposure period: baseline and wave 1. Within each stratum, the stratifying variable was omitted from the adjustment set because it was constant.*

**Supplementary Table S34.** Pre-test of the parallel-trends assumption for the complete-case analysis within subgroups under the upper-quartile definition (unadjusted models).

| Outcome | Lower education *(p-value)* | Higher education *(p-value)* | Older (≥65 years) *(p-value)* | Younger (<65 years) *(p-value)* |
|---|---|---|---|---|
| Body mass index (kg/m²) | 0.783 | 0.913 | 0.845 | 0.306 |
| Overweight/obesity | 0.487 | 0.748 | 0.433 | 0.505 |
| Total cholesterol (mg/dL) | 0.920 | 0.504 | 0.701 | 0.783 |
| HDL cholesterol (mg/dL) | 0.894 | 0.872 | 0.739 | 0.952 |
| LDL cholesterol (mg/dL) | 0.869 | 0.553 | 0.662 | 0.792 |
| Systolic blood pressure (mmHg) | **0.015**† | 0.146 | **<0.001**† | 0.629 |
| Diastolic blood pressure (mmHg) | 0.068 | 0.155 | 0.229 | 0.418 |
| Current tobacco use | 0.434 | 0.083 | 0.323 | 0.769 |
| Current alcohol use | 0.559 | 0.841 | 0.792 | 0.094 |
| Nightly sleep duration (hours) | 0.296 | 0.111 | 0.605 | 0.967 |

*Education: lower education (≤9 years, ISCED 0–2), higher education (>12 years, ISCED 5–8). † $p < 0.05$ (bold) indicates a violation of the parallel-trends assumption, in which case the corresponding ATT is not interpretable as a causal effect. Pre-exposure period: baseline and wave 1. Within each stratum, the stratifying variable was omitted from the adjustment set because it was constant.*

**Supplementary Table S35.** Pre-test of the parallel-trends assumption for the complete-case analysis within subgroups under the upper-quartile definition (adjusted models).

| Outcome | Lower education *(p-value)* | Higher education *(p-value)* | Older (≥65 years) *(p-value)* | Younger (<65 years) *(p-value)* |
|---|---|---|---|---|
| Body mass index (kg/m²) | 0.952 | 0.904 | 0.749 | 0.774 |
| Overweight/obesity | 0.549 | 0.486 | 0.543 | 0.884 |
| Total cholesterol (mg/dL) | 0.751 | 0.721 | 0.691 | 0.537 |
| HDL cholesterol (mg/dL) | 0.944 | 0.673 | 0.764 | 0.886 |
| LDL cholesterol (mg/dL) | 0.627 | 0.318 | 0.513 | 0.498 |
| Systolic blood pressure (mmHg) | 0.060 | **0.041**† | **0.014**† | **0.028**† |
| Diastolic blood pressure (mmHg) | 0.128 | 0.148 | 0.136 | 0.129 |
| Current tobacco use | 0.505 | 0.766 | 0.549 | 0.846 |
| Current alcohol use | 0.875 | 0.257 | 0.850 | 0.106 |
| Nightly sleep duration (hours) | 0.211 | 0.594 | 0.274 | 0.944 |

*Education: lower education (≤9 years, ISCED 0–2), higher education (>12 years, ISCED 5–8). † $p < 0.05$ (bold) indicates a violation of the parallel-trends assumption, in which case the corresponding ATT is not interpretable as a causal effect. Pre-exposure period: baseline and wave 1. Within each stratum, the stratifying variable was omitted from the adjustment set because it was constant.*

**Supplementary Table S36.** Pre-test of the parallel-trends assumption for the IPCW analysis within subgroups under the upper-quartile definition (unadjusted models).

| Outcome | Lower education *(p-value)* | Higher education *(p-value)* | Older (≥65 years) *(p-value)* | Younger (<65 years) *(p-value)* |
|---|---|---|---|---|
| Body mass index (kg/m²) | 0.981 | 0.880 | 0.746 | 0.330 |
| Overweight/obesity | 0.458 | 0.568 | 0.508 | 0.579 |
| Total cholesterol (mg/dL) | 0.986 | 0.534 | 0.744 | 0.771 |
| HDL cholesterol (mg/dL) | 0.996 | 0.936 | 0.801 | 0.806 |
| LDL cholesterol (mg/dL) | 0.881 | 0.586 | 0.645 | 0.760 |
| Systolic blood pressure (mmHg) | **0.010**† | 0.147 | **<0.001**† | 0.775 |
| Diastolic blood pressure (mmHg) | 0.057 | 0.140 | **0.024**† | 0.655 |
| Current tobacco use | 0.365 | 0.163 | 0.286 | 0.903 |
| Current alcohol use | 0.557 | 0.877 | 0.781 | 0.086 |
| Nightly sleep duration (hours) | 0.208 | 0.090 | 0.464 | 0.976 |

*Education: lower education (≤9 years, ISCED 0–2), higher education (>12 years, ISCED 5–8). † $p < 0.05$ (bold) indicates a violation of the parallel-trends assumption, in which case the corresponding ATT is not interpretable as a causal effect. Pre-exposure period: baseline and wave 1. Within each stratum, the stratifying variable was omitted from the adjustment set because it was constant.*

**Supplementary Table S37.** Pre-test of the parallel-trends assumption for the IPCW analysis within subgroups under the upper-quartile definition (adjusted models).

| Outcome | Lower education *(p-value)* | Higher education *(p-value)* | Older (≥65 years) *(p-value)* | Younger (<65 years) *(p-value)* |
|---|---|---|---|---|
| Body mass index (kg/m²) | 0.990 | 0.385 | 0.666 | 0.795 |
| Overweight/obesity | 0.557 | 0.697 | 0.519 | 0.829 |
| Total cholesterol (mg/dL) | 0.696 | 0.552 | 0.620 | 0.540 |
| HDL cholesterol (mg/dL) | 0.998 | 0.808 | 0.826 | 0.863 |
| LDL cholesterol (mg/dL) | 0.545 | 0.641 | 0.433 | 0.512 |
| Systolic blood pressure (mmHg) | **0.033**† | 0.112 | **0.010**† | 0.306 |
| Diastolic blood pressure (mmHg) | 0.116 | 0.294 | 0.123 | 0.247 |
| Current tobacco use | 0.522 | 0.433 | 0.597 | 0.874 |
| Current alcohol use | 0.858 | 0.183 | 0.853 | 0.106 |
| Nightly sleep duration (hours) | 0.201 | 0.729 | 0.253 | 0.923 |

*Education: lower education (≤9 years, ISCED 0–2), higher education (>12 years, ISCED 5–8). † $p < 0.05$ (bold) indicates a violation of the parallel-trends assumption, in which case the corresponding ATT is not interpretable as a causal effect. Pre-exposure period: baseline and wave 1. Within each stratum, the stratifying variable was omitted from the adjustment set because it was constant.*

**Supplementary Table S38.** Final aggregate ATT within subgroups for the complete-case analysis under the ≥1.5 SD definition (unadjusted models).

| Outcome | Lower education ATT (95% CI) | Higher education ATT (95% CI) | Older (≥65 years) ATT (95% CI) | Younger (<65 years) ATT (95% CI) |
|---|---|---|---|---|
| Body mass index (kg/m²) | −0.496 (−1.591, 0.598) | 0.202 (−1.069, 1.473) | −0.743 (−1.693, 0.208) | 0.379 (−1.000, 1.758) |
| Overweight/obesity | −0.074 (−0.187, 0.039) | 0.073 (−0.113, 0.258) | −0.040 (−0.141, 0.060) | −0.045 (−0.212, 0.123) |
| Total cholesterol (mg/dL) | 1.352 (−15.114, 17.817) | −6.306 (−35.877, 23.265) | −1.955 (−18.155, 14.245) | 14.387 (−6.385, 35.159) |
| HDL cholesterol (mg/dL) | 2.389 (−2.578, 7.357) | 0.641 (−4.016, 5.298) | 2.158 (−1.893, 6.210) | 3.188 (−1.923, 8.299) |
| LDL cholesterol (mg/dL) | 7.152 (−8.359, 22.663) | −2.656 (−27.219, 21.907) | −0.343 (−15.332, 14.645) | 16.430 (−3.763, 36.623) |
| Systolic blood pressure (mmHg) | 4.747 (−0.766, 10.259) | −4.770 (−12.632, 3.092) | 2.262 (−2.691, 7.216) | 2.387 (−5.488, 10.262) |
| Diastolic blood pressure (mmHg) | 2.437 (−0.709, 5.582) | 2.558 (−2.596, 7.712) | 2.076 (−0.774, 4.926) | **4.303 (0.204, 8.402)** |
| Current tobacco use | −0.022 (−0.086, 0.042) | 0.039 (−0.113, 0.191) | −0.039 (−0.088, 0.009) | 0.044 (−0.115, 0.204) |
| Current alcohol use | −0.066 (−0.203, 0.071) | −0.129 (−0.301, 0.043) | −0.057 (−0.188, 0.073) | −0.012 (−0.136, 0.112) |
| Nightly sleep duration (hours) | −0.332 (−0.701, 0.038) | 0.169 (−0.398, 0.736) | **−0.401 (−0.775, −0.028)** | 0.043 (−0.438, 0.525) |

*Education: lower education (≤9 years, ISCED 0–2), higher education (>12 years, ISCED 5–8). ATT, average treatment effect on the treated; CI, confidence interval; Bold denotes statistically significant results. Within each stratum, the stratifying variable was omitted from the adjustment set because it was constant.*

**Supplementary Table S39.** Final aggregate ATT within subgroups for the complete-case analysis under the ≥1.5 SD definition (adjusted models).

| Outcome | Lower education ATT (95% CI) | Higher education ATT (95% CI) | Older (≥65 years) ATT (95% CI) | Younger (<65 years) ATT (95% CI) |
|---|---|---|---|---|
| Body mass index (kg/m²) | −1.077 (−2.432, 0.278) | 0.638 (−0.792, 2.068) | −0.918 (−2.122, 0.285) | 0.400 (−0.735, 1.535) |
| Overweight/obesity | −0.113 (−0.253, 0.026) | −0.043 (−0.257, 0.171) | −0.070 (−0.217, 0.078) | −0.066 (−0.224, 0.092) |
| Total cholesterol (mg/dL) | −19.849 (−48.602, 8.903) | −9.274 (−41.883, 23.335) | −28.197 (−63.298, 6.904) | 13.726 (−0.303, 27.755) |
| HDL cholesterol (mg/dL) | 2.818 (−7.870, 13.506) | 2.416 (−3.612, 8.444) | **−12.016 (−18.983, −5.050)** | 1.361 (−2.833, 5.555) |
| LDL cholesterol (mg/dL) | −14.902 (−37.869, 8.066) | 7.892 (−18.347, 34.131) | −21.004 (−47.066, 5.058) | 13.492 (−2.752, 29.735) |
| Systolic blood pressure (mmHg) | **7.014 (1.437, 12.590)** | −5.617 (−14.203, 2.969) | 4.223 (−0.939, 9.385) | 4.238 (−4.716, 13.192) |
| Diastolic blood pressure (mmHg) | 3.330 (−0.327, 6.987) | 1.394 (−4.271, 7.059) | 2.652 (−0.733, 6.038) | **5.813 (0.847, 10.779)** |
| Current tobacco use | −0.013 (−0.071, 0.045) | 0.112 (−0.076, 0.300) | −0.038 (−0.090, 0.015) | 0.030 (−0.134, 0.194) |
| Current alcohol use | −0.054 (−0.197, 0.090) | −0.157 (−0.348, 0.034) | −0.059 (−0.194, 0.077) | 0.000 (−0.124, 0.124) |
| Nightly sleep duration (hours) | −0.280 (−0.734, 0.173) | 0.284 (−0.353, 0.921) | **−0.373 (−0.685, −0.061)** | −0.034 (−0.507, 0.439) |

*Education: lower education (≤9 years, ISCED 0–2), higher education (>12 years, ISCED 5–8). ATT, average treatment effect on the treated; CI, confidence interval; Bold denotes statistically significant results. Within each stratum, the stratifying variable was omitted from the adjustment set because it was constant.*

**Supplementary Table S40.** Final aggregate ATT within subgroups for the IPCW analysis under the ≥1.5 SD definition (unadjusted models).

| Outcome | Lower education<br>ATT (95% CI) | Higher education<br>ATT (95% CI) | Older (≥65 years)<br>ATT (95% CI) | Younger (<65 years)<br>ATT (95% CI) |
|---|---|---|---|---|
| Body mass index (kg/m²) | −0.772 (−1.950, 0.406) | 0.277 (−1.094, 1.649) | −0.936 (−1.974, 0.102) | 0.409 (−0.893, 1.711) |
| Overweight/obesity | −0.077 (−0.206, 0.051) | 0.067 (−0.124, 0.258) | −0.053 (−0.174, 0.069) | −0.059 (−0.231, 0.114) |
| Total cholesterol (mg/dL) | −1.673 (−18.645, 15.298) | −5.814 (−32.501, 20.872) | −2.820 (−18.228, 12.588) | 18.251 (−3.965, 40.468) |
| HDL cholesterol (mg/dL) | 2.188 (−2.578, 6.953) | 0.457 (−4.395, 5.310) | 1.685 (−1.856, 5.225) | 3.675 (−1.431, 8.781) |
| LDL cholesterol (mg/dL) | 4.919 (−10.220, 20.058) | −3.123 (−27.452, 21.205) | 0.497 (−14.695, 15.689) | **22.949 (6.574, 39.325)** |
| Systolic blood pressure (mmHg) | 5.502 (−0.361, 11.364) | −4.659 (−12.039, 2.721) | 2.935 (−2.357, 8.226) | 2.302 (−5.749, 10.354) |
| Diastolic blood pressure (mmHg) | 2.376 (−1.006, 5.758) | 2.820 (−2.245, 7.885) | 1.998 (−0.900, 4.896) | 4.181 (−0.234, 8.596) |
| Current tobacco use | −0.021 (−0.079, 0.036) | 0.064 (−0.109, 0.237) | −0.039 (−0.089, 0.011) | 0.045 (−0.118, 0.209) |
| Current alcohol use | −0.069 (−0.208, 0.070) | −0.127 (−0.292, 0.038) | −0.059 (−0.187, 0.068) | −0.013 (−0.152, 0.125) |
| Nightly sleep duration (hours) | −0.402 (−0.829, 0.025) | 0.179 (−0.402, 0.760) | **−0.501 (−0.893, −0.109)** | 0.063 (−0.471, 0.598) |

*Education: lower education (≤9 years, ISCED 0–2), higher education (>12 years, ISCED 5–8). ATT, average treatment effect on the treated; CI, confidence interval; Bold denotes statistically significant results. Within each stratum, the stratifying variable was omitted from the adjustment set because it was constant.*

**Supplementary Table S41.** Final aggregate ATT within subgroups for the IPCW analysis under the ≥1.5 SD definition (adjusted models).

| Outcome | Lower education<br>ATT (95% CI) | Higher education<br>ATT (95% CI) | Older (≥65 years)<br>ATT (95% CI) | Younger (<65 years)<br>ATT (95% CI) |
|---|---|---|---|---|
| Body mass index (kg/m²) | −0.888 (−2.082, 0.305) | 0.590 (−0.850, 2.030) | −0.775 (−2.057, 0.507) | 0.432 (−0.669, 1.533) |
| Overweight/obesity | −0.071 (−0.255, 0.113) | −0.036 (−0.249, 0.177) | −0.039 (−0.232, 0.155) | −0.081 (−0.252, 0.090) |
| Total cholesterol (mg/dL) | **−36.163 (−60.443, −11.882)** | −8.610 (−39.909, 22.687) | −39.352 (−78.920, 0.216) | **14.070 (0.401, 27.740)** |
| HDL cholesterol (mg/dL) | 1.966 (−9.304, 13.235) | 2.180 (−3.955, 8.315) | **−12.985 (−20.868, −5.101)** | 1.035 (−2.780, 4.850) |
| LDL cholesterol (mg/dL) | −17.884 (−44.797, 9.029) | 6.944 (−13.385, 27.273) | −24.755 (−57.745, 8.235) | 14.352 (−2.632, 31.336) |
| Systolic blood pressure (mmHg) | **7.624 (1.703, 13.545)** | −5.127 (−13.980, 3.726) | 4.634 (−0.533, 9.802) | 4.101 (−5.009, 13.211) |
| Diastolic blood pressure (mmHg) | 3.586 (−0.004, 7.176) | 1.522 (−4.018, 7.062) | 2.763 (−0.797, 6.322) | 4.065 (−0.651, 8.781) |
| Current tobacco use | −0.020 (−0.078, 0.039) | 0.098 (−0.091, 0.287) | −0.042 (−0.092, 0.008) | 0.044 (−0.139, 0.226) |
| Current alcohol use | −0.054 (−0.186, 0.078) | −0.142 (−0.37, 0.086) | −0.059 (−0.195, 0.076) | −0.004 (−0.125, 0.117) |
| Nightly sleep duration (hours) | −0.394 (−0.854, 0.067) | 0.251 (−0.401, 0.903) | **−0.543 (−1.048, −0.038)** | −0.039 (−0.541, 0.463) |

*Education: lower education (≤9 years, ISCED 0–2), higher education (>12 years, ISCED 5–8). ATT, average treatment effect on the treated. CI, confidence interval; Bold denotes statistically significant results. Within each stratum, the stratifying variable was omitted from the adjustment set because it was constant.*

**Supplementary Table S42.** Final aggregate ATT within subgroups for the complete-case analysis under the upper-quartile definition (unadjusted models).

| Outcome | Lower education<br>ATT (95% CI) | Higher education<br>ATT (95% CI) | Older (≥65 years)<br>ATT (95% CI) | Younger (<65 years)<br>ATT (95% CI) |
|---|---|---|---|---|
| Body mass index (kg/m²) | 0.485 (−0.482, 1.452) | 0.011 (−0.893, 0.914) | −0.022 (−0.778, 0.734) | 0.428 (−0.427, 1.283) |
| Overweight/obesity | 0.039 (−0.057, 0.134) | 0.067 (−0.049, 0.183) | 0.015 (−0.067, 0.097) | 0.041 (−0.075, 0.157) |
| Total cholesterol (mg/dL) | −3.710 (−15.351, 7.931) | −0.294 (−15.086, 14.498) | −5.627 (−16.534, 5.281) | −0.649 (−13.003, 11.705) |
| HDL cholesterol (mg/dL) | −0.788 (−3.935, 2.360) | 2.764 (−1.148, 6.677) | 0.600 (−1.892, 3.092) | 0.126 (−3.393, 3.645) |
| LDL cholesterol (mg/dL) | −0.353 (−11.246, 10.540) | 0.269 (−12.409, 12.947) | −5.344 (−15.954, 5.267) | 1.521 (−9.482, 12.524) |
| Systolic blood pressure (mmHg) | 1.874 (−3.089, 6.838) | 4.522 (−2.163, 11.207) | 1.406 (−2.499, 5.311) | **8.803 (2.151, 15.454)** |
| Diastolic blood pressure (mmHg) | 0.896 (−1.731, 3.522) | **4.560 (0.384, 8.736)** | 1.579 (−0.672, 3.829) | 3.964 (−0.682, 8.609) |
| Current tobacco use | 0.051 (−0.006, 0.108) | 0.033 (−0.060, 0.127) | 0.006 (−0.038, 0.051) | 0.047 (−0.059, 0.154) |
| Current alcohol use | −0.006 (−0.134, 0.121) | 0.060 (−0.068, 0.188) | 0.049 (−0.051, 0.149) | 0.035 (−0.069, 0.139) |
| Nightly sleep duration (hours) | −0.309 (−0.674, 0.056) | −0.035 (−0.402, 0.332) | −0.288 (−0.583, 0.006) | −0.306 (−0.688, 0.077) |

*Education: lower education (≤9 years, ISCED 0–2), higher education (>12 years, ISCED 5–8). ATT, average treatment effect on the treated; CI, confidence interval; Bold denotes statistically significant results. Within each stratum, the stratifying variable was omitted from the adjustment set because it was constant.*

**Supplementary Table S43.** Final aggregate ATT within subgroups for the complete-case analysis under the upper-quartile definition (adjusted models).

| Outcome | Lower education<br>ATT (95% CI) | Higher education<br>ATT (95% CI) | Older (≥65 years)<br>ATT (95% CI) | Younger (<65 years)<br>ATT (95% CI) |
|---|---|---|---|---|
| Body mass index (kg/m²) | 0.520 (−0.366, 1.405) | −0.531 (−1.598, 0.536) | 0.332 (−0.476, 1.140) | 0.394 (−0.477, 1.264) |
| Overweight/obesity | 0.045 (−0.065, 0.155) | 0.072 (−0.069, 0.213) | 0.060 (−0.044, 0.165) | 0.025 (−0.085, 0.134) |
| Total cholesterol (mg/dL) | −9.133 (−27.671, 9.404) | 2.847 (−13.921, 19.615) | −18.259 (−38.953, 2.435) | 1.015 (−11.889, 13.919) |
| HDL cholesterol (mg/dL) | −0.317 (−4.562, 3.927) | 4.213 (−0.512, 8.938) | 2.588 (−1.879, 7.056) | −0.310 (−3.879, 3.258) |
| LDL cholesterol (mg/dL) | −6.219 (−22.275, 9.837) | −7.164 (−21.938, 7.610) | −16.582 (−33.504, 0.339) | 3.997 (−8.538, 16.532) |
| Systolic blood pressure (mmHg) | 2.224 (−2.502, 6.951) | 5.937 (−1.418, 13.292) | 1.853 (−2.466, 6.171) | 7.115 (−0.213, 14.443) |
| Diastolic blood pressure (mmHg) | 0.228 (−2.572, 3.028) | 3.617 (−1.098, 8.332) | 0.937 (−1.605, 3.479) | 2.941 (−1.783, 7.665) |
| Current tobacco use | 0.036 (−0.018, 0.091) | −0.024 (−0.137, 0.089) | 0.028 (−0.018, 0.074) | 0.053 (−0.054, 0.160) |
| Current alcohol use | 0.014 (−0.107, 0.136) | 0.083 (−0.058, 0.224) | 0.052 (−0.059, 0.163) | 0.017 (−0.084, 0.117) |
| Nightly sleep duration (hours) | −0.176 (−0.598, 0.245) | −0.102 (−0.389, 0.185) | −0.243 (−0.598, 0.113) | −0.268 (−0.643, 0.108) |

*Education: lower education (≤9 years, ISCED 0–2), higher education (>12 years, ISCED 5–8). ATT, average treatment effect on the treated; CI, confidence interval; Within each stratum, the stratifying variable was omitted from the adjustment set because it was constant.*

**Supplementary Table S44.** Final aggregate ATT within subgroups for the IPCW analysis under the upper-quartile definition (unadjusted models).

| Outcome | Lower education<br>ATT (95% CI) | Higher education<br>ATT (95% CI) | Older (≥65 years)<br>ATT (95% CI) | Younger (<65 years)<br>ATT (95% CI) |
|---|---|---|---|---|
| Body mass index (kg/m²) | 0.405 (−0.513, 1.323) | −0.233 (−1.204, 0.739) | 0.117 (−0.674, 0.909) | 0.237 (−0.684, 1.157) |
| Overweight/obesity | 0.048 (−0.069, 0.164) | 0.038 (−0.093, 0.169) | 0.038 (−0.058, 0.134) | 0.001 (−0.119, 0.121) |
| Total cholesterol (mg/dL) | −11.351 (−26.150, 3.447) | −2.451 (−17.496, 12.594) | −13.520 (−27.516, 0.476) | 0.535 (−12.387, 13.457) |
| HDL cholesterol (mg/dL) | −1.984 (−5.224, 1.255) | 3.901 (−0.374, 8.176) | −0.621 (−3.741, 2.499) | 0.904 (−2.990, 4.799) |
| LDL cholesterol (mg/dL) | −6.980 (−19.726, 5.766) | −4.054 (−17.949, 9.840) | −11.675 (−23.860, 0.511) | 1.944 (−11.252, 15.139) |
| Systolic blood pressure (mmHg) | 1.276 (−3.833, 6.386) | 4.681 (−2.264, 11.626) | 0.861 (−3.080, 4.801) | **8.532 (2.073, 14.991)** |
| Diastolic blood pressure (mmHg) | 0.418 (−2.511, 3.348) | **4.484 (0.322, 8.646)** | 1.137 (−1.312, 3.587) | 3.503 (−0.820, 7.827) |
| Current tobacco use | 0.051 (−0.007, 0.109) | 0.031 (−0.075, 0.136) | 0.017 (−0.031, 0.065) | 0.041 (−0.067, 0.150) |
| Current alcohol use | −0.008 (−0.127, 0.110) | 0.062 (−0.075, 0.198) | 0.045 (−0.059, 0.150) | 0.041 (−0.061, 0.142) |
| Nightly sleep duration (hours) | **−0.418 (−0.819, −0.016)** | −0.094 (−0.462, 0.274) | **−0.424 (−0.775, −0.073)** | −0.336 (−0.705, 0.032) |

*Education: lower education (≤9 years, ISCED 0–2), higher education (>12 years, ISCED 5–8). ATT, average treatment effect on the treated; CI, confidence interval; Bold denotes statistically significant results. Within each stratum, the stratifying variable was omitted from the adjustment set because it was constant.*

**Supplementary Table S45.** Final aggregate ATT within subgroups for the IPCW analysis under the upper-quartile definition (adjusted models).

| Outcome | Lower education<br>ATT (95% CI) | Higher education<br>ATT (95% CI) | Older (≥65 years)<br>ATT (95% CI) | Younger (<65 years)<br>ATT (95% CI) |
|---|---|---|---|---|
| Body mass index (kg/m²) | 0.628 (−0.258, 1.514) | 0.412 (−0.503, 1.327) | 0.514 (−0.313, 1.341) | 0.330 (−0.590, 1.250) |
| Overweight/obesity | 0.079 (−0.044, 0.203) | 0.061 (−0.068, 0.190) | 0.093 (−0.029, 0.215) | 0.012 (−0.118, 0.141) |
| Total cholesterol (mg/dL) | −19.630 (−43.782, 4.523) | −8.174 (−34.926, 18.578) | −24.842 (−51.927, 2.244) | 2.891 (−11.328, 17.110) |
| HDL cholesterol (mg/dL) | −1.759 (−7.302, 3.784) | 1.284 (−4.019, 6.587) | 0.737 (−4.371, 5.844) | 0.045 (−3.577, 3.668) |
| LDL cholesterol (mg/dL) | −15.863 (−36.242, 4.516) | −6.539 (−28.147, 15.069) | −23.633 (−47.516, 0.251) | 5.423 (−8.902, 19.749) |
| Systolic blood pressure (mmHg) | 2.369 (−2.786, 7.524) | 3.128 (−2.317, 8.573) | 1.795 (−2.720, 6.311) | 2.647 (−4.183, 9.477) |
| Diastolic blood pressure (mmHg) | 0.213 (−2.840, 3.265) | 1.417 (−1.732, 4.566) | 0.876 (−1.645, 3.397) | 1.938 (−2.104, 5.980) |
| Current tobacco use | 0.042 (−0.013, 0.097) | 0.028 (−0.031, 0.087) | 0.037 (−0.013, 0.087) | 0.052 (−0.054, 0.159) |
| Current alcohol use | 0.013 (−0.112, 0.139) | 0.034 (−0.093, 0.161) | 0.050 (−0.059, 0.158) | 0.019 (−0.078, 0.116) |
| Nightly sleep duration (hours) | −0.326 (−0.803, 0.152) | −0.241 (−0.738, 0.256) | −0.382 (−0.820, 0.057) | −0.305 (−0.720, 0.110) |

*Education: lower education (≤9 years, ISCED 0–2), higher education (>12 years, ISCED 5–8). ATT, average treatment effect on the treated; CI, confidence interval; Within each stratum, the stratifying variable was omitted from the adjustment set because it was constant.*

### Section G. Sensitivity analysis including participants who moved between differently classified census block groups

Forty participants moved between census block groups with different gentrification classifications during the post-exposure period (27 from a gentrifying to a non-gentrifying census block group and 13 in the opposite direction) and were excluded from the primary analysis. In this sensitivity analysis, they were retained and classified according to their census block group of residence at wave 3, the first post-exposure wave analysed. IPCW weights were re-estimated in the expanded sample using the same specification as in the primary analysis.

**Supplementary Table S46.** Final aggregate ATT and pre-test of the parallel-trends assumption for the IPCW analysis including participants who moved between differently classified census block groups (adjusted and unadjusted models).

| Outcome | Adjusted model | | Unadjusted model | |
|---|---|---|---|---|
| | ATT (95% CI) | Pre-test (p) | ATT (95% CI) | Pre-test (p) |
| Body mass index (kg/m²) | −0.081 (−0.798, 0.637) | 0.997 | −0.006 (−0.688, 0.676) | 0.914 |
| Overweight/obesity | 0.006 (−0.088, 0.100) | 0.268 | 0.007 (−0.068, 0.082) | 0.214 |
| Current tobacco use | 0.009 (−0.033, 0.052) | 0.023† | 0.030 (−0.013, 0.074) | 0.046† |
| Current alcohol use | −0.050 (−0.128, 0.028) | 0.319 | −0.046 (−0.118, 0.025) | 0.782 |
| Total cholesterol (mg/dL) | −8.211 (−21.491, 5.068) | 0.050† | 1.860 (−7.372, 11.091) | 0.154 |
| HDL cholesterol (mg/dL) | −0.250 (−4.602, 4.101) | 0.305 | 1.095 (−1.434, 3.624) | 0.360 |
| LDL cholesterol (mg/dL) | −4.758 (−16.488, 6.973) | 0.014† | 2.202 (−6.398, 10.802) | 0.064 |
| Systolic blood pressure (mmHg) | 1.824 (−2.188, 5.836) | 0.010† | 1.447 (−2.406, 5.300) | <0.001† |
| Diastolic blood pressure (mmHg) | 1.024 (−1.379, 3.426) | 0.882 | 0.920 (−1.165, 3.004) | 0.302 |
| Nightly sleep duration (hours) | −0.316 (−0.605, −0.027) | 0.183 | −0.408 (−0.661, −0.155) | 0.523 |

*ATT, average treatment effect on the treated; CI, confidence interval; IPCW, inverse probability of censoring weighting. Participants who moved between differently classified census block groups during the post-exposure period were classified according to their census block group of residence at wave 3. Models adjusted for sex, age, education, marital status, and employment. 95% CIs derived from the influence function. † $p < 0.05$ indicates a violation of the parallel-trends assumption, in which case the corresponding ATT is not interpretable as a causal effect (unrounded $p = 0.0499$ for total cholesterol, adjusted model). Pre-exposure period: baseline and wave 1.*
